# Understanding patterns of variant emergence and spread in an ongoing epidemic

**DOI:** 10.64898/2026.03.27.26349560

**Authors:** Anjalika Nande, Michael Z. Levy, Alison L. Hill

## Abstract

The COVID-19 pandemic saw successive emergence and global spread of novel viral variants, exhibiting enhanced transmissibility or evasion of immunity. While the genotypic and phenotypic basis of SARS-CoV-2 variants have been extensively characterized, the evolutionary factors governing their patterns of emergence are less well understood. In this study we systematically investigated how the invasion dynamics of viral variants depend on variant phenotype (increased transmissibility or immune evasion), source (local evolution vs importation), the timing of introduction, the distribution of population susceptibility, and the contact network structure. Using a stochastic multi-strain epidemic model, we find that strains with only a transmission advantage are more likely to emerge earlier in the epidemic, and rapidly and predictably dominate the viral population. In contrast, immune-escape variants tend to linger at low prevalence for extended time periods after emergence, avoiding detection, until a critical amount of immunity has built up in the population and they begin to rapidly outcompete existing strains. We find that two common features of realistic human contact networks—heterogeneity in contacts (overdispersion) and clustering—lead to more punctuated evolutionary dynamics. This work provides insight into past dynamics of SARS-CoV-2 variants and can help define planning scenarios for future epidemic modeling efforts.

## Introduction

A defining feature of the COVID-19 pandemic has been the repeated emergence of variants of concern that have compromised public health measures and contributed to successive waves of disease over several years [1]. SARS-CoV-2 variants like Alpha (B.1.1.7 [2]), Delta (B.1.617.2 [3]), and Omicron (B.1.1.529 [4]) were first detected in specific locations before eventually spreading globally and completely replacing existing strains. While the genetic basis of these variants and their phenotypic effects on cellular infectivity, antibody neutralization, transmissibility and vaccine efficacy have been extensively characterized [5], much less is known about the underlying evolutionary mechanisms that drive their dynamics.

Substantial research, both theoretical and empirical, has been focused on understanding other aspects of the evolutionary dynamics of infectious disease: the long-term coexistence of diverse strains [6–8], the process of evolutionary rescue facilitating spillover into a new host species [9, 10], and the evolution of drug resistance during therapy [11–13]. However, less attention has been paid to the early-time dynamics of emerging pathogen variants during the epidemic phase of a disease. What factors contributed to the extremely rapid replacement of resident SARS-CoV-2 strains by each successive variant? Is the timing of variant detection and takeover primarily determined by the time of introduction or the stage of the epidemic in the population? When do we expect to see variants that primarily confer a transmission advantage vs those that primarily evade existing immune responses? When using models for scenario planning during epidemics, what scenarios for variant emergence should be considered? Are any aspects of variant emergence predictable?

Existing studies of early variant dynamics have tended to focus on quantifying the selective advantage of emerging variants compared to circulating strains. Many different models have been developed for this purpose [14–19], often producing different estimates. Existing estimation methods are also often sensitive to location-specific factors like population structure and levels of immunity that affect the speed of variant spread. For example, Earnest et. al [20] estimated that the SARS-CoV-2 Delta variant has a 63%–167% selective advantage over the Alpha variant (range of averages across US New England states), and Van Dorp et. al [16] estimated a 42%–125% selective advantage of the Omicron variant over Delta (range of medians across 40 countries). To add to this complexity, early dynamics of variants are subject to stochastic effects due to small population sizes right after emergence, which can obscure true selective advantages; stochasticity has been shown to accelerate early epidemic growth and substantially influence initial trajectories, complicating efforts to characterize emerging variants [21]. Moreover, identifying the nature of this advantage—whether the variant has selective advantage due to its immune evasive properties or due to a higher transmission rate or some combination of the two—is also important as it affects variant impact on mitigation measures and epidemic planning scenarios. However, different combinations of transmission advantage and immune evasion can result in the same overall selective advantage, making it impossible to identify the relative contributions of each solely using data on variant frequencies over time [22]. Identifying differences in variant emergence patterns depending upon their type could help in disentangling these contributions and improve estimation methods. Most importantly, methods that focus solely on quantifying the dynamics of observable variants cannot contribute to our understanding of the likelihood of variant emergence.

In this study we systematically evaluate the factors that determine the probability of variant invasion during outbreaks, the speed at which new variants spread and displace existing strains, and the overall impact of variant emergence on the epidemic trajectory. Using a simplified evo-epidemiological model, we examined how variant phenotype (increased transmissibility or immune evasion), source (local evolution vs importation), timing of introduction, distribution of population susceptibility, and the contact network structure (e.g. highly over-dispersed, clustered) interact to determine the fate of new variants. Inspired by SARS-CoV-2 variants of concern but designed to give general insight, we believe this approach is a necessary step towards developing better methods to interpret real-world data on variant frequencies and relate such data to underlying phenotypic effects. Moreover, we hope to provide intuition for when different types of variants are most likely to be detected during ongoing epidemics.

## Methods

### Model

We developed a simplified stochastic SIR-type model of a resident and a variant strain to study the dynamics of variant invasion and spread (Figure 1). Individuals in the population are classified as susceptible to infection (*S*); infected with the resident (*I_r_*) or the variant (*I_v_*) strain; or recovered from infection with the resident (*R_r_*) or the variant strain (*R_v_*). The resident and variant strain have different per contact rates of transmission to susceptible individuals (*β_r_* and *β_v_*, respectively). We define the transmission advantage as *σ* = *β_v_/β_r_*. The infectious period can follow an arbitrary distribution with average duration 1*/γ_i_* for strain *i*, which here we assume to be equal for both strains. Recovered individuals are assumed to be immune to re-infection in the absence of immune escape variants. We model immune evasion by defining an escape parameter ε (0 ≤ ε ≤ 1) which determines the efficiency at which the variant can infect individuals with immunity to the resident strain. When ε = 0, the variant has no immune evasive properties and can not infect individuals with immunity to the resident strain (i.e., in *R_r_*). When *ε* = 1, the variant achieves 100% immune evasion and can infect these previously-infected individuals as if they were fully susceptible. In the Supplement, we consider model extensions, such as allowing immunity to re-infection with the same strain to wane on same the time-scale as the initial epidemic peak and variant emergence.

**Figure 1:**
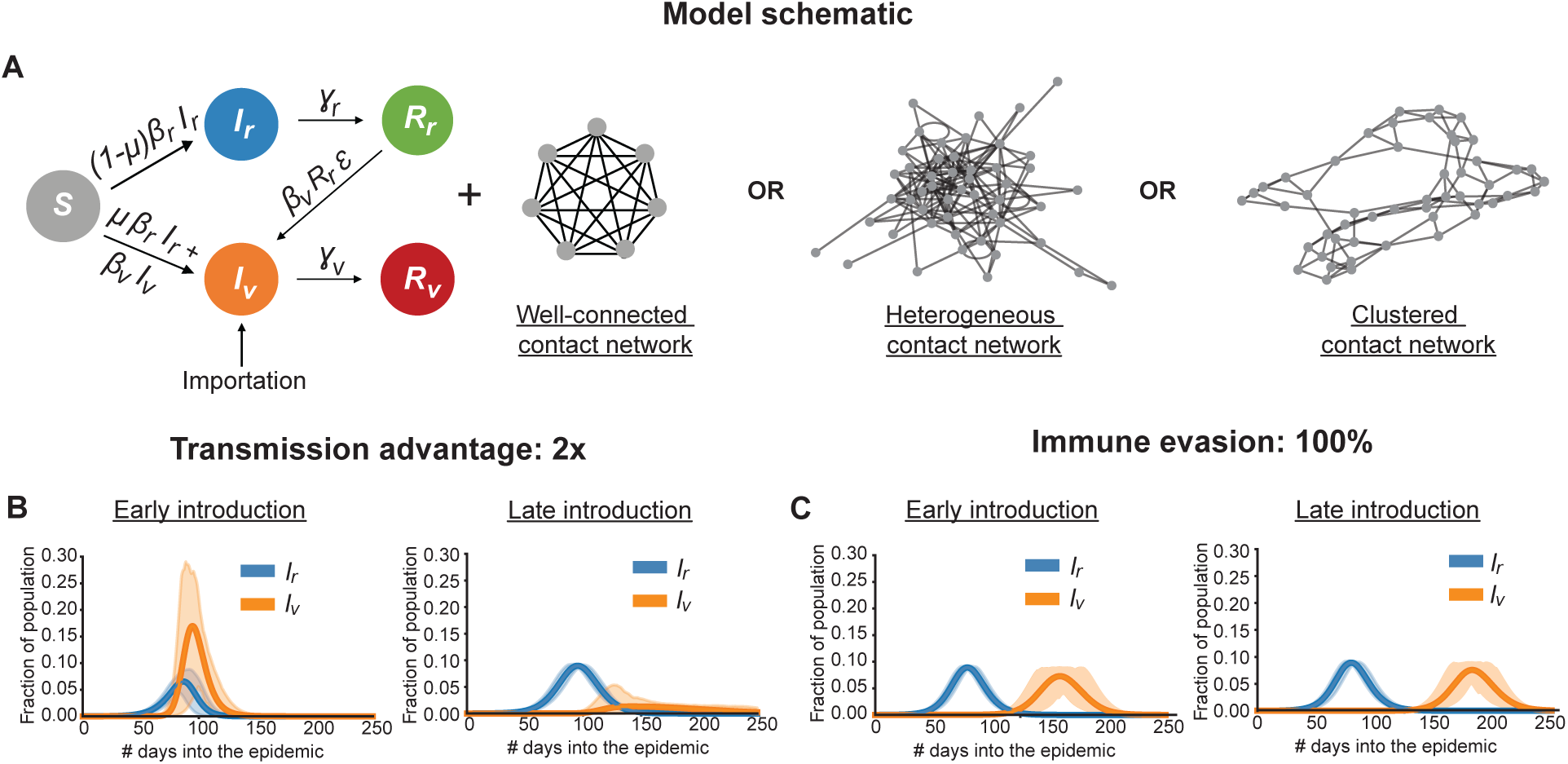
Model schematic and example infection dynamics. A) Schematic of the two strain SIR-type model with the different types of contact networks considered consisting of individuals susceptible to infection (*S*), infected by the resident (*I_r_*) or variant (*I_v_*) strain, and recovered from infection by the resident (*R_r_*) or variant (*R_v_*) strain. For strain i, *β_i_* is the per contact transmission rate and *γ_i_* is the rate of recovery. The variant can be produced via mutation (rate *µ*) of the resident strain or imported into the population from an outside source. Variants with immune evasive properties (0 *< ε <*= 1) can infect individuals recovered from the resident strain infection by a rate proportional to the strength of immune evasion *ε*. Example infection dynamics for a variant with B) transmission advantage and with C) immune evasive properties when introduced early or late in the epidemic. Results show the mean and 95% CI over 100 iterations.

We consider two modes of variant introduction: importation, where at pre-specified times in the simulation a variant infection is introduced into the population (e.g., from an outbreak in another population), and local evolution, where each infection with the resident strain can lead to transmission of a variant infection (i.e., due to within-host evolution) with a fixed probability *µ*.

We simulate infection spreading stochastically over a fixed contact network. For our main analysis, we consider a well-connected population where individuals can potentially infect anyone else. Later, we compare these results with those from more realistic contact patterns that take into account the effects of heterogeneity in the number of contacts and clustering of immunity (Figures 1A).

Similar to most models of infectious disease spread, the fitness of each strain can be encapsulated by the basic reproductive number, *R*_0_, which is defined as the average number of new infections generated by one infected individual introduced in a population of fully susceptible individuals, and its real-time analogue *R_t_*. For our model parameters, *R*_0_ for the two strains and *R_t_* for the variant at the time of its introduction *t_int_* obtained from their respective single strain models is given by,

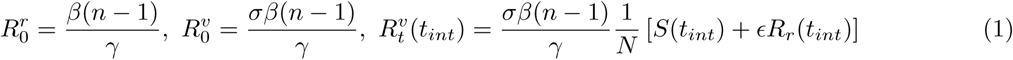

where *N* and *n* are the total population size and the mean degree of the network respectively. As a baseline, we assume 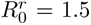 and that both the strains have the same gamma-distributed duration of the infectious period (7 ± 4 days). The per contact transmission rate *β* is then calculated via Equation 1 for given *n, σ* and *ε*. Example dynamics of this model are provided in the Figures 1, S1, and S2. These values were chosen to approximately match the infectious period of SARS-CoV-2 [23], but the qualitative results we obtain are unaffected by this choice of parameter values.

Our stochastic model is simulated as a discrete time stochastic process and is implemented in the Python package JAX Numpy which is optimized to run simulations on GPUs, allowing us to simulate the whole epidemic in a few minutes even for population sizes of a million. Our model and implementation code is open access and available on GitHub: https://github.com/anjalika-nande/variant-emergence-patterns.

### Simulation details

#### Calculating the invasion probability of the variant

We first seed a resident strain infection (*I_r_*(*t* = 0) = 20) into a fully susceptible population of size 10,000. There are two ways in which a variant can then be introduced into the simulation at pre-specified times: a susceptible individual acquires a variant infection from outside of the population (importation), or the variant evolves in an individual infected with the resident strain (local evolution). We assume variant importation for most of the results presented here, but also consider local evolution. The introduction method does not affect the subsequent variant invasion dynamics in well-mixed populations, but in more structured contact networks the clustering of infected individuals can result in different invasion probabilities for variants that emerge via local evolution vs via importation. We choose variant importation times based on the cumulative size of the resident strain epidemic at the time of introduction. This allows us to study the impact of population immunity on variant invasion. We consider a range of epidemic sizes, from 0.5% – 50% of the population infected by the resident strain at the time of variant introduction.

To calculate the invasion probability of the variant, we run multiple simulations (10, 000 replicates) of our stochastic model with the mutation probability set to zero and for each iteration introduce one individual infected by the variant at the desired stage of the simulation. The simulation is run until zero infected individuals of either type remain. The invasion probability is then calculated as the fraction of iterations where the variant avoided early stochastic extinction. Practically, we classify an invasion as successful if the final epidemic size of the variant reaches at least 100 infections, a threshold we found reliably distinguishes sustained spread from early stochastic extinction and is independent of overall population size. Note that our invasion probability is *not* analogous to a measure of fixation probability in traditional population genetics models—there is no real concept of fixation here as both disease strains eventually go extinct as susceptibility wanes and the epidemic runs its course. Thus we choose an arbitrary (but we believe biologically relevant) threshold to define invasion, and for example, our invasion probabilities for neutral variants will not correspond to long-term expectations based on genetic drift.

#### Calculating variant prevalence over time

To investigate the dynamics of invading variants, we track prevalence of the variant as a function of time since introduction for simulations in which the variant successfully invaded. These sets of simulations were run for a larger population size of 1 million to represent a typical metropolitan area. Note, this larger population size was not used for the invasion probability simulations since running 10,000 replicates would have been computationally too time consuming and would not have altered the invasion probability. Prevalence is defined as the fraction of infections at any given time that are due to the variant (e.g., *I_v_/*(*I_r_* + *I_v_*)). The variant is said to have invaded if it cumulatively infects at least 100 individuals and prevalence is averaged only for these successful invasions. We analyze trajectories of 50 successful invasions for each set of parameter values. For each replicate of the simulation we calculate the time for the variant to reach 5%, 50%, and 95% prevalence after introduction. We call the time at which the variant reaches 5% the ‘detection’ time and the time taken for the variant to sweep from 5% – 95% prevalence the ‘takeover’ time.

### Simulating variant evolution

To simulate local evolution of the variant, we assume that, each new infection caused by an individual infected with the resident strain has a probability *µ* of instead being a variant infection. This is an over-simplification as we are encapsulating all the potentially relevant processes underlying the creation of the variant—such as, evolution through step-wise mutations or evolution in chronically infected individuals—in a single parameter. But this assumption is valid for our purposes to investigate the population-level invasion dynamics of variants produced via within-host de novo evolution in infected individuals since it captures the effects of more infections leading to a higher chance of variant production, and a faster rate of evolution leading to an earlier introduction of the variant.

We define ‘low’, ‘intermediate’ and ‘high’ rates of evolution depending upon the number of resident strain infections needed on average before 1 variant is produced. A low rate corresponds to one variant infection produced, on average, after 1 million resident strain infections, an intermediate rate after 10^5^ infections, and a high rate after 10^4^ infections. This translates to *µ* equaling 1*e*^−6^, 1*e*^−5^ and 1*e*^−4^ for low to high respectively. For reference, in the United Kingdom, the Alpha variant of SARS-CoV-2 was detected after ∼ 1.3 × 10^4^ cumulative confirmed cases of the ancestral strain per million individuals [24, 25].

### Details of contact networks

#### Well-connected network

Following [62], we approximate a well-mixed population by randomly connecting each person in the population to 100 other individuals. Although an ideal “well-mixed” network would involve connecting every individual to everyone else, given the large population sizes that we use (up to 1 million), this would require an enormous amount of computational memory and compromise the efficiency of using sparse matrices to represent the networks. Considering that in reality individuals typically transmit infections to only a limited number of others before recovering, a uniformly random network with a high degree serves as a good approximation of a fully-connected network.

### Network with heterogeneity in the number of contacts

To study how heterogeneity in the number of individual contacts affects variant invasion and spread, we construct a network where the number of contacts individuals have follows a negative binomial distribution. There is no further structure in the network and individuals are randomly connected to each other. For baseline parameters we choose the coefficient of variation *CV* = 0.7 and the mean network degree *µ_network_* = 15 and back out the parameters of the negative binomial distribution using the following relationships,

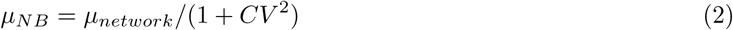

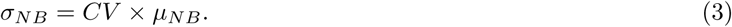

We choose the mean network degree to be 15 to roughly reflect the average number of daily contacts individuals have reported in studies conducted in US [63] and in European [64] populations. The coefficient of variation was chosen to get sufficient spread in the number of contacts.

### Network with clustering of contacts

To study the effect of clustering of infections and consequently of immunity on variant invasion and spread, we construct a network where there is a high chance that the contacts of an individual are also contacts of each other. One way of creating such clustered networks is by using the Watts-Strogatz small-world algorithm [32]. We use the NetworkX python package [66] implementation of this method via the connected_watts_strogatz_graph function to create the network structure. We assume that all individuals have the same number of contacts which is again set to 15 to roughly reflect the average number of contacts per person observed in the US. We choose 0.2 as the probability of rewiring as for a network with mean degree of 15 this produces a network with a clustering coefficient ∼ 0.3 which is the order of magnitude of clustering observed among human social contacts [29, 67].

## Results

### Invasion probability and epidemic impact

We first investigate how likely a new variant is to establish an outbreak depending upon its phenotype (enhanced transmissibility vs immune evasion) and what stage of the epidemic it first appears (Figures 1B, 2). We focus on two metrics of variant success: the fraction of simulations where the variant avoids early stochastic extinction, which is a proxy for the invasion probability, and the size of the variant outbreak relative to the whole outbreak (to gauge the overall impact of the variant on the epidemic). A neutral variant (identical to the resident strain) serves as a baseline. We note that the trends discussed here are unaffected by the specific choices of parameter values (see Suppl. Figures S3, S4, S5).

We find that for variants with a transmission advantage, the chance of invasion (versus extinction) increases with earlier introduction and with larger transmission advantage (Figure 2A). If introduced in the very early stages of an epidemic, even a neutral variant can have a significant chance of invasion purely on account of drift [26], and strains with even modest transmission advantages are highly likely to invade. For example, for the simulated scenarios, a neutral variant introduced after only 1% of the population had been previously infected had a 32% chance of invasion, compared to 73% for a strain with 1.5x transmission advantage and 92% with 3x transmission advantage. After early invasion, variants with a transmission advantage are likely to dominate the epidemic, increasing peak incidence and cumulative infections (Fig. 1B, 2B,C). If transmission advantage variants appear later in the epidemic, they are only likely to invade if they have large fitness advantages. For example, if they appear when 20% of the population has already been infected, the invasion probability drops to 0.39% for neutral variants, 30% for 1.5x advantage and only slightly—to 90%—for 3x transmission advantage. However, even if the chance of invasion is high, variants introduced later in the epidemic often fail to dominate the outbreak (Figure 2D). Even a highly transmissible variant (3x advantage) introduced after 20% prior infection dominates in only ∼ 32% of simulations.

**Figure 2:**
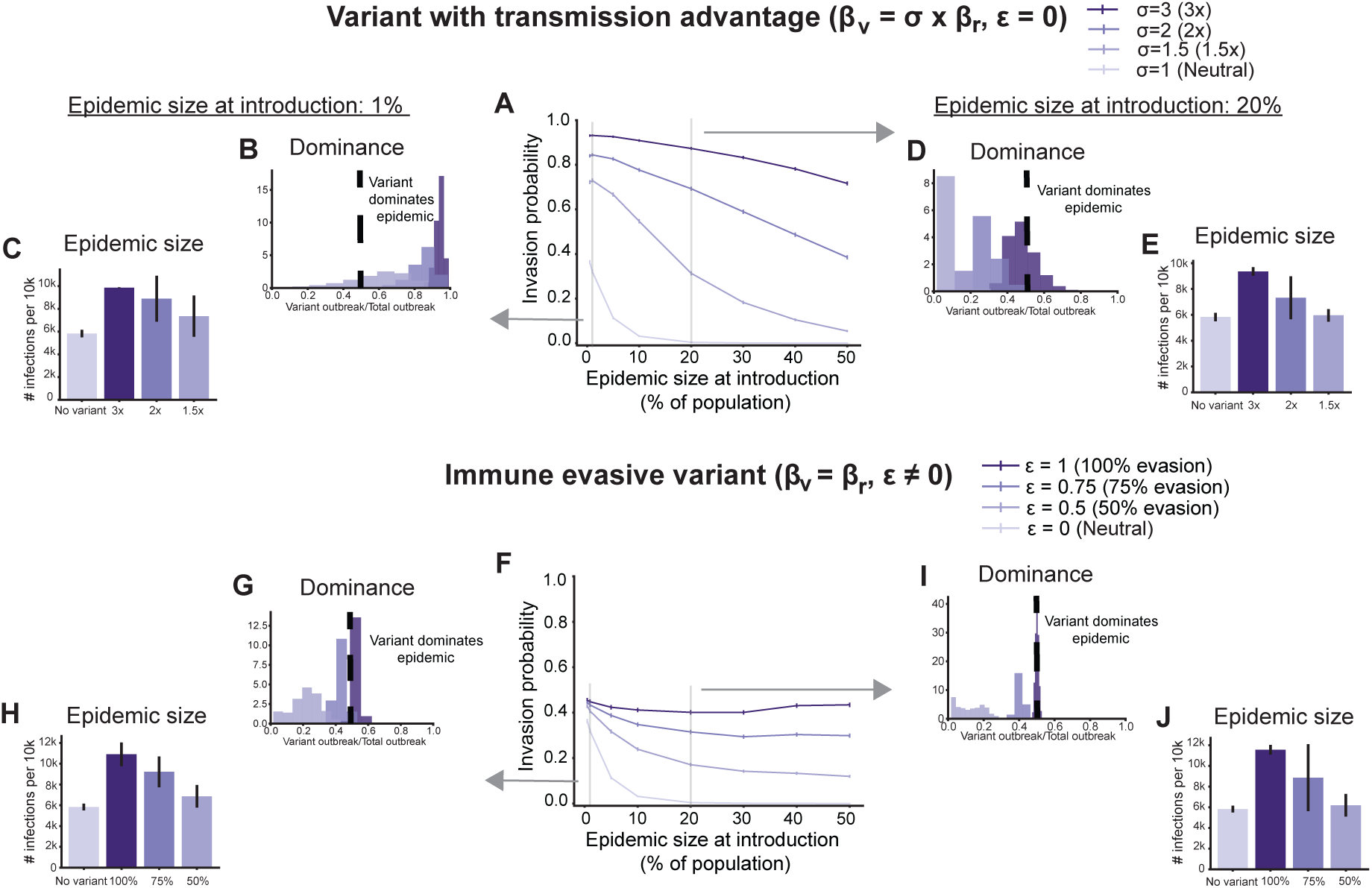
Effect of variant type and population immunity on variant invasion and epidemic impact. Results of stochastic simulations for variants with transmission advantage (top row) and variants with immune evasive properties (bottom row). A)&F) Fraction of simulations where the variant avoided early stochastic extinction as a function of the size of the resident strain epidemic at the time of introduction. The vertical lines are the standard error of proportion. Histograms showing the relative size of the variant outbreak compared to the total epidemic size, and the average total epidemic size conditional on non-extinction, for two introduction times: when the variant is introduced after 1% (B, C, G, H) or 20% (D, E, I, J) of the population has been infected by the resident strain. Vertical black lines on the epidemic size bars correspond to 2 standard deviations on either side of the mean. Darker colors correspond to variants with a higher advantage for both variant types. Results are for 10,000 replicates for each introduction time.

Population immunity at the time of introduction plays a big role in these trends. Although the relative fitness advantage of a variant compared to the resident strain does not depend on the introduction time, the absolute fitness at the time of introduction for a strain with only a transmission advantage is lower in more immune populations (Figure S6A). As a result, even if the transmission advantage of the variant is high, if introduced after a significant proportion of the population has already been infected, it can have only a limited impact. A consequence of this dynamic is that the overall impact of a successful variant on the epidemic—as measured by the increase in the total cumulative infections by the time the epidemic ends (also known as final attack size)—depends mainly on the variant fitness advantage and less so on the introduction time (Figures 2C,E). While variants introduced earlier tend to replace infections with the resident strain, the excess infections are similar and determined mainly by the difference in *R*_0_ between the strains.

For immune escape variants, we find that the chance of invasion increases with earlier introductions and higher levels of immune evasion (Figure 2F). As the epidemic progresses, unless immune escape is complete, immune escape variants have a lower invasion probability (higher probability of extinction) because there are fewer opportunities for secondary infections when prior immunity is higher (Figure S7). Note, however, that in the early epidemic stages when there is little immunity from prior infection, these variants are essentially neutral (Figure S6B) compared to the resident strain—they have the same baseline *R*_0_ and are competing for the exact same susceptible pool—and so the invasion probability is close to that of a neutral variant irrespective of the degree of immune escape. In agreement with previous work [27, 28], we find that although the invasion probability can be substantial, immune escape variants rarely dominate the epidemic (Figures 2G,I). For example, a 50% immune evasive variant introduced after the resident strain has infected 1% of the population accounts for only 22% of total infections on average. This lack of dominance, however, does not imply a small overall epidemic, as depending upon the degree of immune evasion, the size of the variant outbreak can be close to that of the resident strain even if it does not exceed it. As a result, the final epidemic size can still be large and even exceed the population size for highly immune evasive variants due to reinfections (Figures 2H,J). While successfully invading variants with a transmission advantage tend to displace the resident strain, especially if they appear early in the epidemic or have large fitness advantages, immune escape strains cause less interference with the resident strain and instead owe their success to reinfections.

### Rate of increase in variant prevalence and consequences for detection

Next, for variants that successfully invade, we investigated the dynamics of their spread by tracking the fraction of all current infections caused by the variant over time (Figure 3). These variant frequency curves are commonly used to quantify variant fitness advantage from pathogen sequence data [14–17]. We find that variants with a transmission advantage increase in prevalence faster if they are introduced early in an epidemic compared to later (Figure 3A-C). This difference is primarily due to faster growth immediately after introduction. For example, a variant twice as transmissible as the resident strain reaches 5% prevalence in an average of 24 days if introduced when 1% of the population was already infected, but takes 42 days if not introduced until 10% prior infection (purple violins, Figure 3B). In contrast, the time taken to go from 5% –95% prevalence—which we define as the variant ‘takeover time’—is fairly consistent across introduction times (pink violins, Figure 3B). For immune escape variants, we find the opposite trend: the earlier they are introduced, the *longer* they take to reach 5% prevalence (Figures 3E-G). Immune evasive variants tend to linger at a low prevalence for long periods of time until a critical level of immunity has built up in the population due to infections with the resident strain epidemic, at which point the variant can quickly take over (Figure 3G). Note that these dynamics are conditional on non-extinction. Although these variants do not actually take over until much later in the epidemic, essentially washing out the effect of their introduction time, their higher probability of invasion in the early epidemic stages still makes it more advantageous for them to arise very early and spread neutrally with respect to the resident strain until they have enough of an advantage to really start out-competing it.

**Figure 3:**
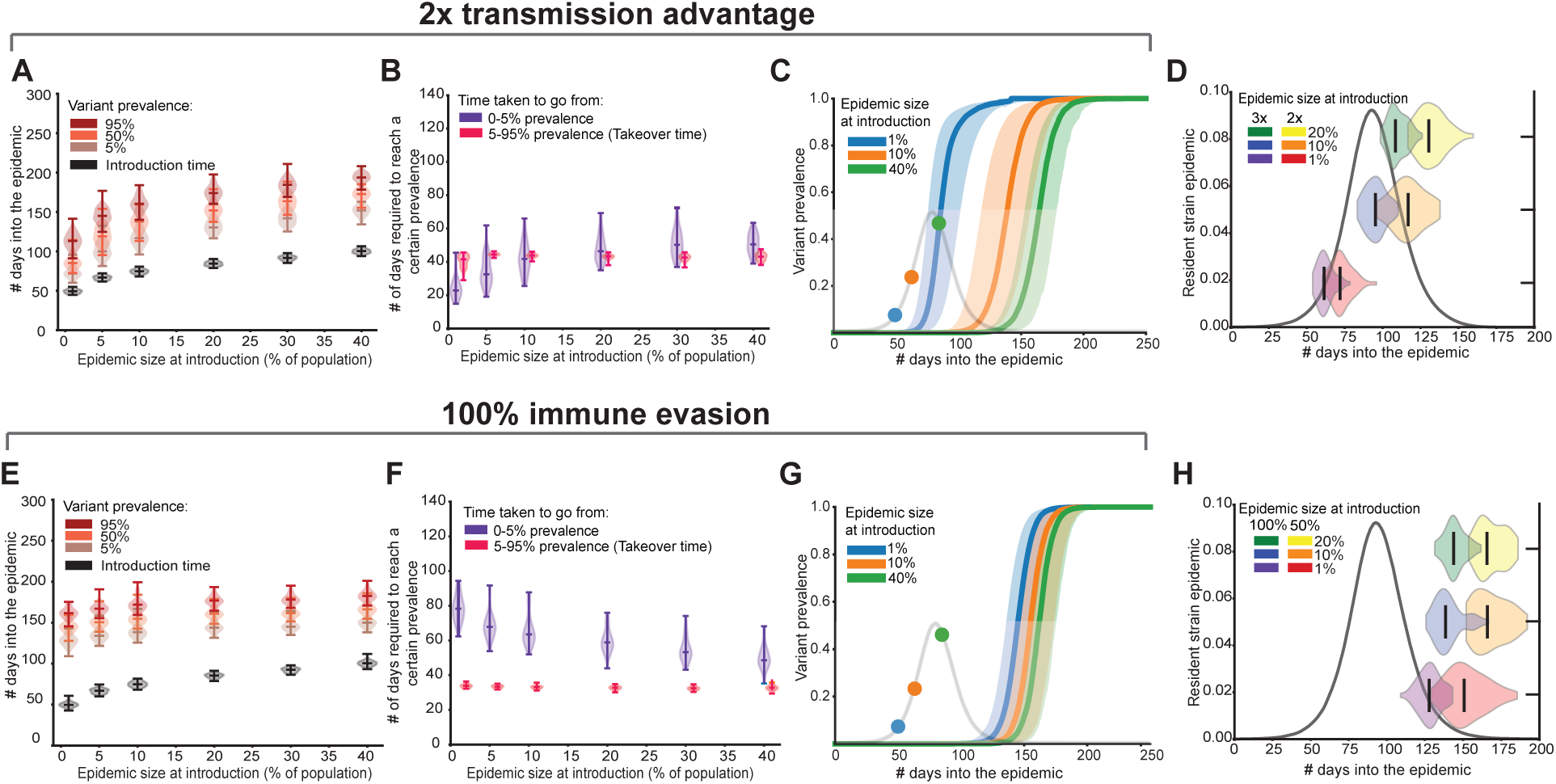
Rate of increase in variant prevalence as a function of the epidemic size at the time of introduction. Example results for a variant with transmission advantage (top row) and for an immune evasive variant (bottom row) when the variant escapes stochastic extinction. A)&E) Time when the variant was introduced (black violins) and reached 5%, 50% and 95% prevalence as a function of the size of the resident strain epidemic at the time of introduction. The violins are marked with the median, min and max values. B)&F) Number of days required for the variant to go from introduction to 5% and 5% – 95% prevalence as a function of the size of the resident strain epidemic at the time of introduction. C)&G) Variant prevalence over time when introduced at different levels of population immunity. Solid line is the median and the shaded region corresponds to the 5-95 percentile range. The resident strain epidemic in the absence of the variant is overlaid in light grey along with the introduction times for reference. D)&H) Violin plots of the time at which the variant reaches 5% prevalence in reference to the resident strain epidemic (black curves) for different introduction times. Results are for 50 iterations for each introduction time in a population of size 1 million.

These trends can be explained by considering the two major regimes in variant invasion dynamics: a stochastic regime immediately after introduction, and a deterministic regime once the variant has escaped stochastic extinction. During the stochastic phase, which coincides roughly with the time from introduction until reaching 5% prevalence, the rate of increase in variant prevalence is influenced by both absolute and relative fitness of the variant. For variants with a transmission advantage, relative fitness is constant over time (Figure S6). Higher absolute fitness drives the observation that variants introduced earlier have a higher probability of invasion and rise in prevalence more rapidly. In contrast, for immune evasive variants, absolute fitness decreases over the course of the epidemic, leading to small decreases in invasion probability, but it is compensated for by increasing relative fitness, leading to faster establishment for later introduction times. Variant dynamics during the 5-95% prevalence sweep (‘takeover’) fall under the deterministic regime, where relative fitness primarily governs the dynamics. Since the relative fitness of variants with a transmission advantage is always constant, the takeover time is unaffected by introduction time. For immune escape variants, although relative fitness increases over time, variants introduced at different times tend to reach 5% prevalence at similar stages in the resident strain epidemic, resulting in similar relative advantages during the takeover phase and thus comparable takeover times.

These differences in variant dynamics have important real-world implications for when different types of variants are likely to be detected during an epidemic. If we approximate ‘detection time’ as the time taken to reach 5% prevalence, our result suggests that variants with a transmission advantage can be detected during any phase of the resident strain epidemic, depending on the timing of introduction and strength of their advantage, but that immune evasive variants, irrespective of their degree of immune evasion or introduction times, will most likely be detected only in the later stages when there is already a significant amount of population immunity (Figures 3D,H, and Supplementary Discussion and Figure S8). These trends are robust to the choice of parameter values (Figures S5, S9, S10)

So far we have modeled variant introduction as a single importation of the variant from an external source. We also considered what happens when variants are generated from current infections with the resident strain with some transition probability, meant to approximate the process of within-host evolution. We find that the trends observed for the timing of variant appearance still hold (Figure 4). For example, even when the rate of variant production is high, a variant that completely escapes immunity is typically detected around 25 days after the epidemic peak, with detection occurring even later for partially immune evasive variants (Figure S11).

**Figure 4:**
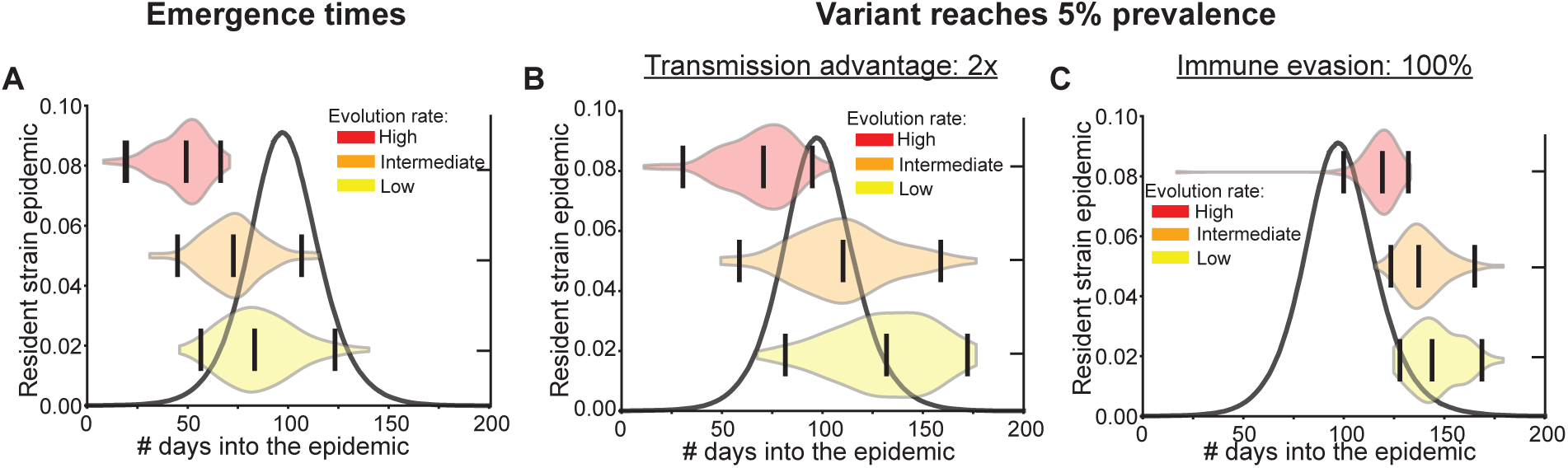
Variant emergence and detection times for different rates of evolution. Time at which variants A) emerge and B)-C) reach 5% prevalence during the resident strain epidemic when the rate of evolution is high (red), intermediate (orange), and low (yellow). The violin plots are marked with the median and the middle 95% quantile range. The resident strain epidemic in the absence of the variant is provided as a reference (black curve). Results are for 100 iterations and population size of a million. Low (10^−6^), medium (10^−5^), and high (10^−4^) evolutionary rates correspond to the emergence of a new variant, on average, after the number of infections equal to the entire population size, 10%, or 1% of the population, respectively. See Methods for more details.

### Effect of contact heterogeneity and clustering on variant dynamics

So far we’ve considered a well-connected population where an infected individual can infect anyone else. This assumption allowed us to isolate the effects of variant type and population immunity on a variant’s ability to invade and spread. However, in reality, human contacts are heterogeneous, they tend to be clustered, and these patterns are known to affect disease transmission in a complicated way [29–34]. To investigate the role played by such realistic human contact patterns on variant invasion dynamics, we repeat the previous analysis on two types of structured populations: a contact network where there is a high degree of heterogeneity in the number of individual contacts, and one where each individual has the same number of contacts but they are clustered, that is, there is a high chance that the contacts of an individual are also contacts of each other (see Methods for more details). This choice allows us to separately study the role played by contact heterogeneity and clustering of immunity on variant invasion dynamics.

We first investigated the effects of heterogeneity and clustering on the variant invasion probability (Figure 5, Figure S12). Across all network types, we observe the same trends as for the well-connected case— irrespective of variant type, invasion becomes less likely later in the resident strain epidemic as the availability of susceptible individuals decreases. However, compared to the well-connected scenario, variants generally find it harder to invade in heterogeneous and clustered networks; with the exception being that for variants with a transmission advantage, clustering of immunity can lower or increase the chance of invasion depending upon the population immunity at the time of emergence and the strength of the transmission advantage.

**Figure 5:**
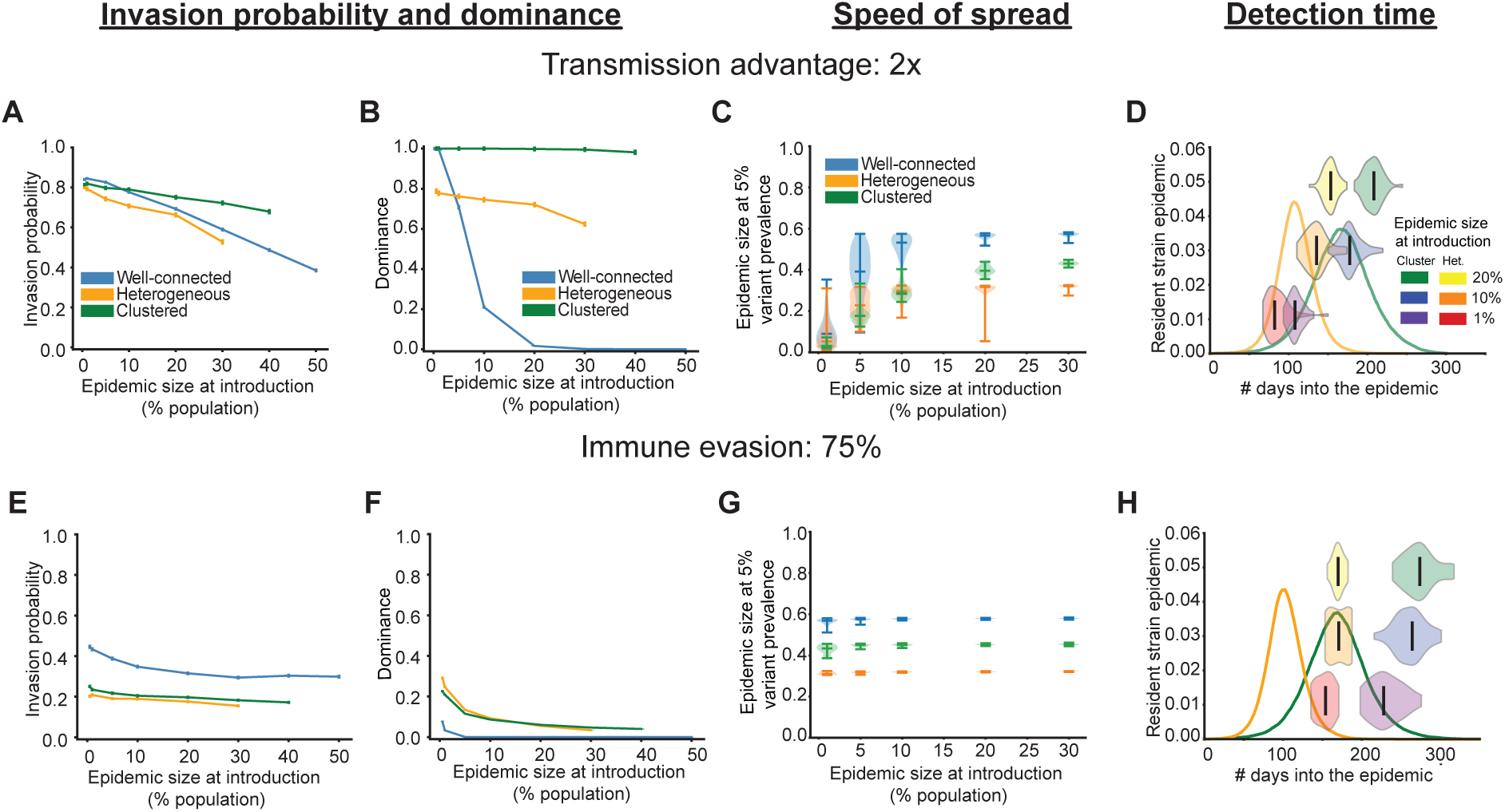
Effect of contact heterogeneity and clustering on variant invasion and spread. Example results for variants with a transmission advantage (top row) and with immune evasive properties (bottom row) when the transmission network is well-connected (blue), heterogeneous (orange) and clustered (green). A)&E) Fraction of simulations when the variant infected more than 1% of the population, and B)&F) fraction of invading runs when the variant dominated the epidemic as a function of the size of the resident strain epidemic at introduction. Vertical marks are the standard error in proportion and results are for ∼ 10^4^ iterations. C)&G) Epidemic size when the variant reaches 5% prevalence versus the epidemic size when it was introduced. D)&H) Violin plots of the time at which the variant reaches 5% prevalence in reference to the resident strain epidemic for the heterogeneous (orange curve) and clustered networks (green curve). Solid middle line is the median and results are for ∼ 50 iterations. See Methods for more details.

A key difference between structured and well-mixed populations is that, in structured populations, the chance of invasion depends on where in the network the variant emerges; for example, a variant is less likely to invade when it is introduced in an individual with fewer contacts as compared to a well-connected one. Consequently, heterogeneity in contact patterns generally lowers the average invasion probability compared to the well-connected case. This also means that the source of the variant—whether imported or locally evolved—matters. When we assume the variant evolves locally, instead of being imported, we find similar qualitative trends (Figures S13, S14). However, for variants with a transmission advantage, the invasion probability is lower under local emergence, since they are more likely to arise in parts of the network with fewer susceptible individuals.

Next, we examine the dynamics of variants that successfully invade. Compared to the well-connected case, heterogeneity and clustering can help variants dominate the epidemic (Figures 5B,F) by helping them spread through the population at a faster rate (Figures 5C,G). Variants that successfully invade usually emerge in the more connected and susceptible parts of the network which allows them to out-compete the resident strain more easily than in the well-connected case. One way to quantify this is by comparing the variant’s rise in prevalence relative to the overall epidemic progression. For example, when a variant twice as transmissible as the resident strain is introduced into a population with 10% prior infection, it reaches 5% prevalence when median cumulative infections are ∼ 50% in the well-connected case, but only ∼30% in structured networks. As a result, the variant dominates the epidemic 100% and 80% of the time in clustered and heterogeneous networks respectively, versus just 20% in the well-connected case. However, although variants increase in prevalence faster, for each network type, our earlier finding that compared to variants with a transmission advantage (Figures 5D and S15A,B), immune evasive variants are more likely to be detected when there are higher levels of population immunity, still holds (Figures 5H and S15C,D).

## Discussion

Continual pathogen evolution is one of the main barriers to infectious disease control, with adaptations commonly leading to escape from immune responses. The COVID-19 pandemic is a recent example of this, where despite vaccines being developed at record-breaking speeds, the successive emergence and dominance of novel viral variants evading immunity has led to a continual burden of disease [35]. Recurring annual outbreaks of seasonal influenza are sustained by similar dynamics, where antigenic evolution occurs both via accumulation of de novo mutations and through genomic reassortment in animal reservoirs [36,37]. Similarly, sustained transmission and sporadic outbreaks of norovirus, particularly the dominant GII.4 strain, is made possible by recurrent emergence of new variants [38, 39]. Although more difficult to measure, viral variants with increased transmissibility have also contributed to emergent or recurrent burden of many infections (e.g. for SARS-CoV-2 [40, 41], influenza [42], West Nile virus [43], Ebola [44]).

Despite innovations in our ability to track pathogen genomic evolution in real-time [45] and measure strain-specific immunity using a high-throughput methods [46–50], the COVID-19 pandemic accentuated the many open questions about the evolutionary dynamics of novel viral variants. What factors contributed to how quickly successive variants emerged from 2021 onwards? Are there differences in the expected likelihood, timing, and dynamics of emergence of variants with a transmission advantage versus immune escape properties? Does superspreading help or hinder variant emergence? Do we expect variant emergence to be synchronized across geographic regions with different infection histories? Understanding all these factors is essential to design epidemic control strategies that can appropriately respond to the ever changing pathogen landscape.

In this study, we use a stochastic epidemic model to examine the invasion dynamics of variants with either transmission advantage or immune evasive properties, in the presence of varying levels of population immunity. We find that, irrespective of the underlying transmission network structure, variants with a transmission advantage or partial immune evasion are more likely to invade and cause large outbreaks when they first appear early in an epidemic when the number of susceptible hosts is high, as they are in direct competition for hosts with existing strains. In contrast, highly immune evasive variants always have access to a largely susceptible population, and their chance of invasion and outbreak size is insensitive to timing of first appearance. While the invasion of immune escape variants can significantly increase epidemic size, these strains are less likely to dominant and outcompete resident strains unless they completely evade existing immunity.

The dynamics of successfully invading variants can be roughly divided into two phases: an initial stochastic phase, where the variant avoids extinction, and a subsequent deterministic one. Our results suggest that the invasion dynamics of the two variant types differ predominantly in the stochastic phase. Variants with a transmission advantage benefit from low levels of population immunity and escape this phase faster the earlier they are introduced in the epidemic, with the rate increasing for increasing levels of advantage. In contrast, successfully invading immune-escape variants increase in prevalence at a much slower rate in the early epidemic stages irrespective of the strength of immune evasion. These variants persist at low prevalence for extended periods, spreading nearly neutrally relative to existing strains until they gain a sufficient relative advantage due to increasing levels of population immunity to existing strains. This suggests that immune evasive variants may evade detection until the later stages of an epidemic. Indeed this is one possible explanation for why the SARS-CoV-2 Omicron variant was detected much later than when phylodynamic analysis suggests it first evolved [51]. Similar patterns have been observed for norovirus variants [52]. As the stochastic phase of invasion typically precedes detection, improving identifiability of the nature of variant fitness advantages will likely require integrating diverse data sources that inform this early phase, such as phylodynamic estimates of introduction times or human mobility-informed models of viral importation [53, 54]. Ultimately, all such approaches would benefit from earlier and more sensitive detection of variant introductions, which could be facilitated by enhanced surveillance systems such as wastewater monitoring [55, 56] or targeted sampling at points of entry (e.g., airports [57, 58]).

To date, most characterization of variant invasion dynamics has focused on measuring the relative instantaneous growth rate (or effective time-varying reproduction number, *R_t_*) of the emerging strain based on case incidence data once the variant has reached a detectable frequency, often using popular software packages (e.g., EpiEstim R package [15, 69]). While such methods will certainly be limited by not informing the probability of invasion or the early stochastic phase of emergence, we hypothesized they are also limited by not explicitly including inter-strain competition for susceptible individuals when estimating variant advantage. Employing EpiEstim on simulation results for transmission-advantage variants (Suppl. Analysis), we found that this approach generally leads to an overestimation of the variant advantage, but occasionally led to underestimation. For situations where the variant was weakly advantageous (e.g. ∼ 1.5x transmissibility) and was introduced into an established epidemic (*>* 10% infected) the error in the estimated advantage was only a few percent (Fig.S16). However, the error was more pronounced for higher fitness variants, earlier introduction times, and increased levels of mixing in the population, with errors up to 25%.

Two prior studies examined the evolutionary epidemiology of emerging variants inspired by SARS-CoV-2 [27, 28]. A key difference of our work is that by using a stochastic model, we are able to examine the invasion probability, timing, and early dynamics, which prior work could not. Examining only the more deterministic phase of variant spread, Bushman *et al* [27] also found that immune escape variants rarely dominate the epidemic, though transmission advantage mutants often do. Reynes *et al* [28] also found the important role of timing of emergence on the subsequent impact of immune escape variants, and found effects suggestive of delayed emergence in some cases.

Our study has several limitations. We haven’t yet considered variants that have *both* a transmission advantage and immune evasive properties, since our focus was on disentangle the differences between the two types of selective advantages. At least in the case of SARS-CoV-2, many variants were suspected to have both advantages, and prior models have included this [27], though it’s also possible that an emerging strain with an improved value of one trait may pay a fitness cost in the other trait. Further work is needed to understand the dynamics of such variants. We don’t include the effects of waning immunity in our model, but our preliminary exploration (and a related deterministic model [28]) suggests that the qualitative trends of invasion observed here are unchanged, and we still find that immune evasive variants are most likely to be detected after the epidemic peaks (see Suppl. Analysis and Figure S17). However, the long-term interaction between waning immunity and immune evasiveness in endemic settings is complicated [59], and left for future work. We also haven’t considered how interventions such as vaccines may affect variants, particularly immune evasive strains (see, e.g. [27, 28]), and especially if there are different population subgroups with different vaccine coverage. While this study focuses on the population-level emergence and selection of variants, these strains must first be generated and selected within individual hosts. Within-host selection is affected by factors not considered here, such as the duration of infection, the relationship between transmissibility and pathogen load, and the dynamics of innate and adaptive immune responses [60, 61]. Finally, we have yet to formally compare the predictions of our models to the extensive spatio-temporal data on SARS-CoV-2 variants, or related data for other pathogens.

In conclusion, our results highlight the epidemiological and evolutionary factors influencing the invasion potential of new pathogen variants during an ongoing outbreak. They shed light on dynamics that drove (and continue to drive) the global burden of COVID-19 and other rapidly-evolving infections, and generate hypotheses that can be tested with pathogen genomic surveillance data. Incorporating the dynamics of variant emergence processes into models, alongside improved surveillance, will enable more accurate risk assessments and, in turn, better inform public health policies.

## Acknowledgments

We thank Thayer Anderson, Madeleine Gastonguay, Anne Hebert, Vivek Murali, members of the Johns Hopkins Infectious Disease Dynamics Group, as well as attendees at EPIDEMICS9, EPIDEMICS10, 32nd Annual Dynamics & Evolution of Human Viruses Conference, and the 2023 Contagion on Complex Social Systems Workshop for helpful feedback.

## Data availability

No data was acquired for this study. All code used to generate the results presented here is open source and available on Github: https://github.com/anjalika-nande/variant-emergence-patterns.

## Supplementary Analysis

### Understanding the extended low prevalence of immune evasive variants after emergence

One of the main results in this work is that immune evasive variants linger at a low prevalence for extended periods of time after emergence and that this is especially pronounced if they emerge in the early stages of the epidemic (Figure 3). We find that this is a consequence of the stochastic nature of variant invasion coupled with the fact that immune evasive variants spread neutrally with respect to the existing strains in the early epidemic stages.

For simplicity, consider a fully immune evasive variant since its dynamics are unaffected by existing strains and it is always exposed to a fully susceptible population at the time of invasion. The time taken for it to avoid stochastic extinction can be roughly calculated by approximating the initial stage of invasion as a constant birth-death process of the population of individuals infected by the variant *I_v_*. The per capita death rate is the rate at which infected individuals recover (*D* = *γ_v_*) and assuming that the depletion of susceptibles at the very early stages of the epidemic is negligible compared to the total population size *N*, the per capita birth rate is given by *B* = *β_v_N* . For such a birth-death process, there exists an analytical solution for the probability of not having gone extinct by a certain time *T* after introduction at time *t* [71],

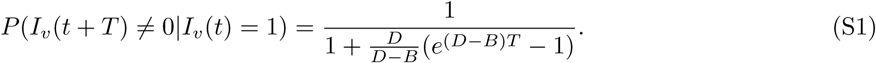

In the limit that *T* → ≃ this gives the establishment probability, which is the probability of avoiding stochastic extinction. We can calculate this existence probability as a function of *T* and if it reaches the establishment probability at a finite time it means that survival is guaranteed past that point and the dynamics can be thought of deterministically. We find that the time taken to avoid stochastic extinction is comparable to the time taken from that point onward to reach the epidemic peak; for example, in a population of 1 million, the time taken for a fully immune evasive variant with 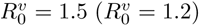 to avoid extinction (defined as reaching within 1% of the establishment probability) is around 60 (160) days according to this approximation whereas, it takes around 80 (120) days on average for the variant epidemic to peak from this point onwards in our simulations. As the baseline *R*_0_ of the variant and the existing strain are the same, this means that as long as the variant emerges when the existing strain is in the deterministic phase, by the time it avoids stochastic extinction and starts increasing faster in prevalence, the existing strain is close to or has passed its epidemic peak (Fig.S8). Note, a fully immune evasive variant escapes stochastic extinction the fastest so this effect is further increased for partially immune evasive variants and they will be detected even later in the epidemic.

### Extension of the model to include effects of waning immunity and seasonality

The dynamics of many human viral infections like influenza and respiratory syncytial virus (RSV) are driven by oscillations in the effective reproductive number of the pathogen due to environmental (for e.g., temperature and humidity) and epidemiological (for e.g., waning of immunity) reasons leading to well-defined yearly peaks in infection [72–75]. To study variant invasion dynamics for such endemic diseases, we extend the previous model to include the effects of waning immunity and seasonality (Figure S17A). We include a waning parameter *ω* which is the rate by which recovered individuals become susceptible to infection, and incorporate the time dependence of environmental factors by allowing the transmission rates (*β*) of the two strains to be time-varying. We assume that this time dependence is sinusoidal [75] and parameterize it such that the effective reproductive number of strain i takes the functional form,

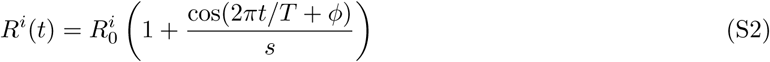

where, *T* is the period of seasonality (which is 1 year by definition), *φ* is the phase of the oscillation and controls when the seasonal oscillation peaks compared to the start of the epidemic (for e.g., *φ*=0 implies the epidemic starts at peak seasonality), and *s* controls the deviation around the basic reproductive number as a result of seasonality. For the scenario considered in Figure S17, the population size is 1 million and we use the same values for 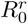 and duration of infectiousness as in the baseline model used in the main text, but choose the duration of recovery to be gamma-distributed (4 ± 1 months) for both the strains and pick *φ* = *π/*2 and *s* = 3. These parameters were chosen as they lead to a well-defined yearly peak in infections (black curve in Figures S17B-D).

We find that for high and intermediate rates of evolution, the detection time results for this model are similar to the baseline as variant emergence occurs early in the epidemic before the effects of seasonality and waning of immunity kick in. However, when the rate of evolution is very low (∼ 1 variant produced on average after 1 million infections which is our total population size), although most emergence still occurs during the first epidemic peak, there is a chance of variant emergence in the inter-season period or during the next peak. Consequently, detection can occur during the inter-season period but it is still most likely after the resident strain epidemic peaks.

### Consequences for estimating variant advantage

To calculate the relative advantage of the variant over the resident strain, we used the estimate_advantage function provided in the R package EpiEstim [15, 69]. The two main inputs required by this function are the time series of the daily incidence data of the two strains and the serial interval distribution. We acquire the daily incidence data directly from our model outputs and calculate the serial interval distribution from the infectious period distribution used in our model via the standard formula [76, 77],

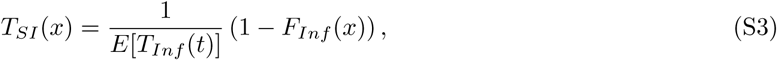

where *T_SI_* is the serial interval distribution, *F_Inf_* is the CDF of the infectious period distribution and *E*[*T_Inf_*] is the expectation value of the infecious period distribution. Note, this formula is generally for the generation time distribution, but as there is no incubation period in the SIR model, the serial interval distribution is the same as the generation time distribution. We used a 7 day sliding window and default values for priors and other MCMC control parameters provided in EpiEstim and validated our approach by ensuring that the resident strain *R_t_* was estimated correctly in the absence of the variant. The final reported value is the mean over ∼ 50 iterations and we used incidence data from the start of the resident strain epidemic until the peak of the variant epidemic. As a sensitivity analysis we varied some of these MCMC parameters and used different time periods of the incidence data, and found that it didn’t change the results qualitatively.

We also computed the relative advantage of the variant using a simpler approach for comparison. To do this we estimate the mean *R_t_* of each of the strains using the relative_R function in EpiEstim. The final relative advantage (solid green line in Figs.S16A,B) is the ratio of the mean *R_t_* (over 50 iterations) of the variant over that of the resident strain and the shaded region is the standard deviation of the distribution of the ratio of the means. We note that the initial higher levels of overestimation and uncertainty seen in these figures is probably a consequence of stochasticity. The equivalent deterministic model trajectory for the system is an average over all the stochastic possibilities, so when only considering runs where invasion occurs, the growth rate of the variant on average is higher than what is expected of a deterministic model (i.e., what is expected based on its baseline *R*_0_) to compensate for the cases where invasion doesn’t occur [21]. We found that the estimation works well when the growth of the variant doesn’t significantly affect the dynamics of the resident strain, that is, when the two strains are essentially undergoing separate epidemics. In situations where this is not true, for example, when the variant has a high transmission advantage or it is introduced in the early epidemic stages, its spread reduces the growth rate of the resident strain due to a faster depletion of the susceptible individuals. Since this reason for the reduction in the growth rate is not accounted for in the estimation, *R_t_* of the resident strain is underestimated, leading to an overestimation of the variant *R_t_* and hence, its relative advantage. This is also seen when comparing across network types— there is less overestimation (and sometimes even underestimation) of the variant advantage when a variant emerges in a highly clustered network. This is because, for a variant to successfully invade it has to emerge in the part of the network that has more susceptible individuals, which is generally not the same part of the network where the resident strain is spreading, and hence, it has a lesser impact on the resident strain *R_t_*.

## Supplementary Figures

**Figure S1:**
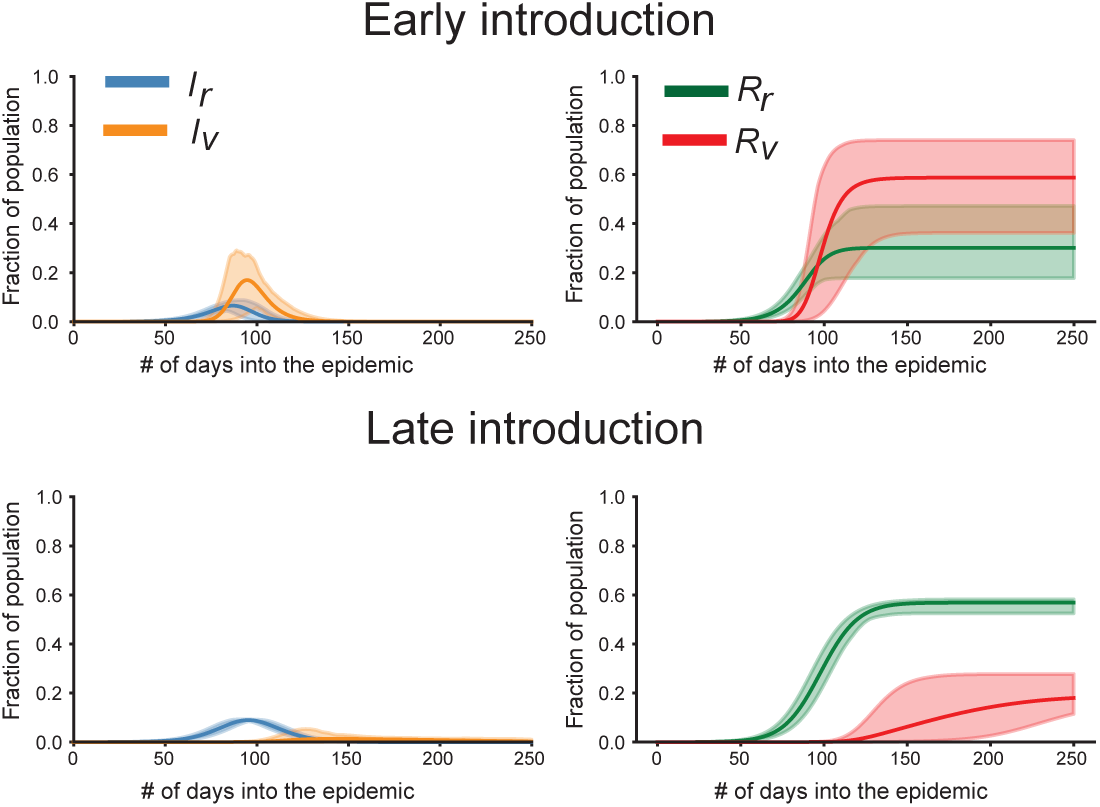
Example dynamics of the two strain model with a transmission advantage variant. Time-course of the number of infected (left) and recovered for the two strains (right) when the variant is introduced early (top row) and late (bottom) in the resident strain epidemic. Early (late) introduction corresponds to the variant introduced when the resident strain has infected 1% (10%) of the population. Results are for resident strain 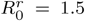 and a variant that is twice as transmissible 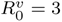. The solid line and shaded region corresponding to the mean and the 5th-95th percentile calculated from 50 iterations of the simulation.

**Figure S2:**
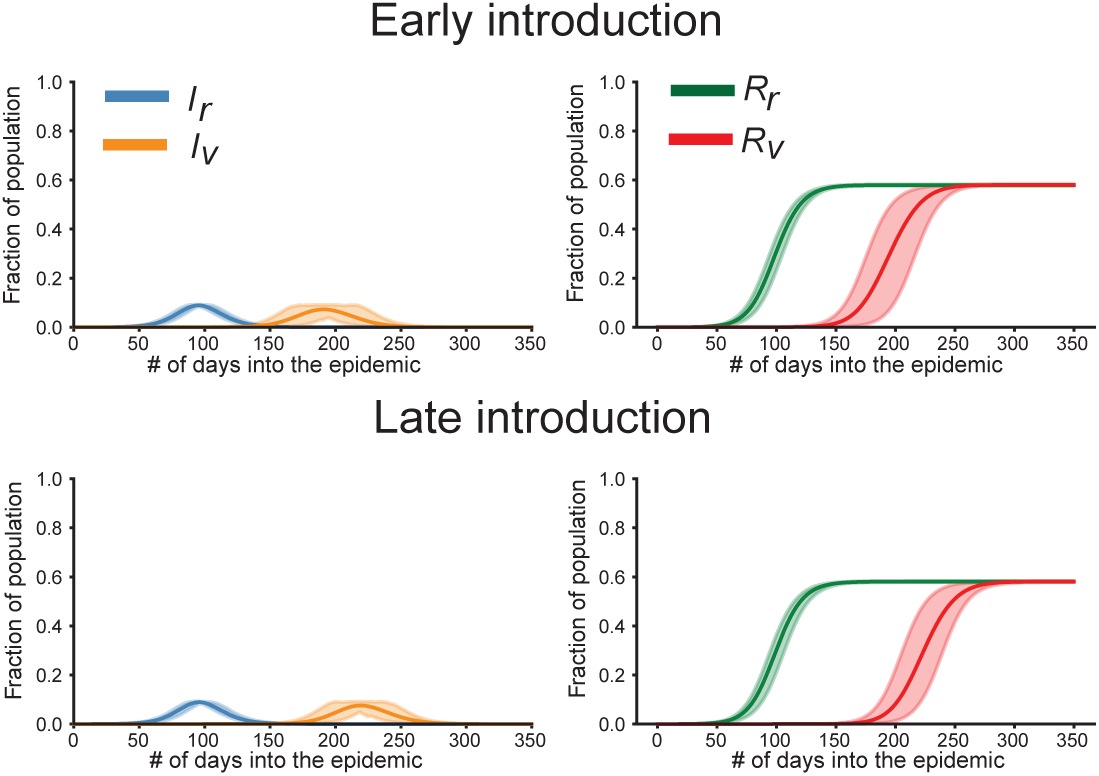
Example dynamics of the two strain model with a immune evasive variant. Time-course of the number of infected (left) and recovered for the two strains (right) when the variant is introduced early (top row) and late (bottom) in the resident strain epidemic. Early (late) introduction corresponds to the variant introduced when the resident strain has infected 1% (20%) of the population. Results are for resident strain 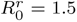 and a fully immune evasion variant (*ε* = 1) variant with no transmission advantage 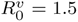. The solid line and shaded region corresponding to the mean and the 5th-95th percentile calculated from 50 iterations of the simulation.

**Figure S3:**
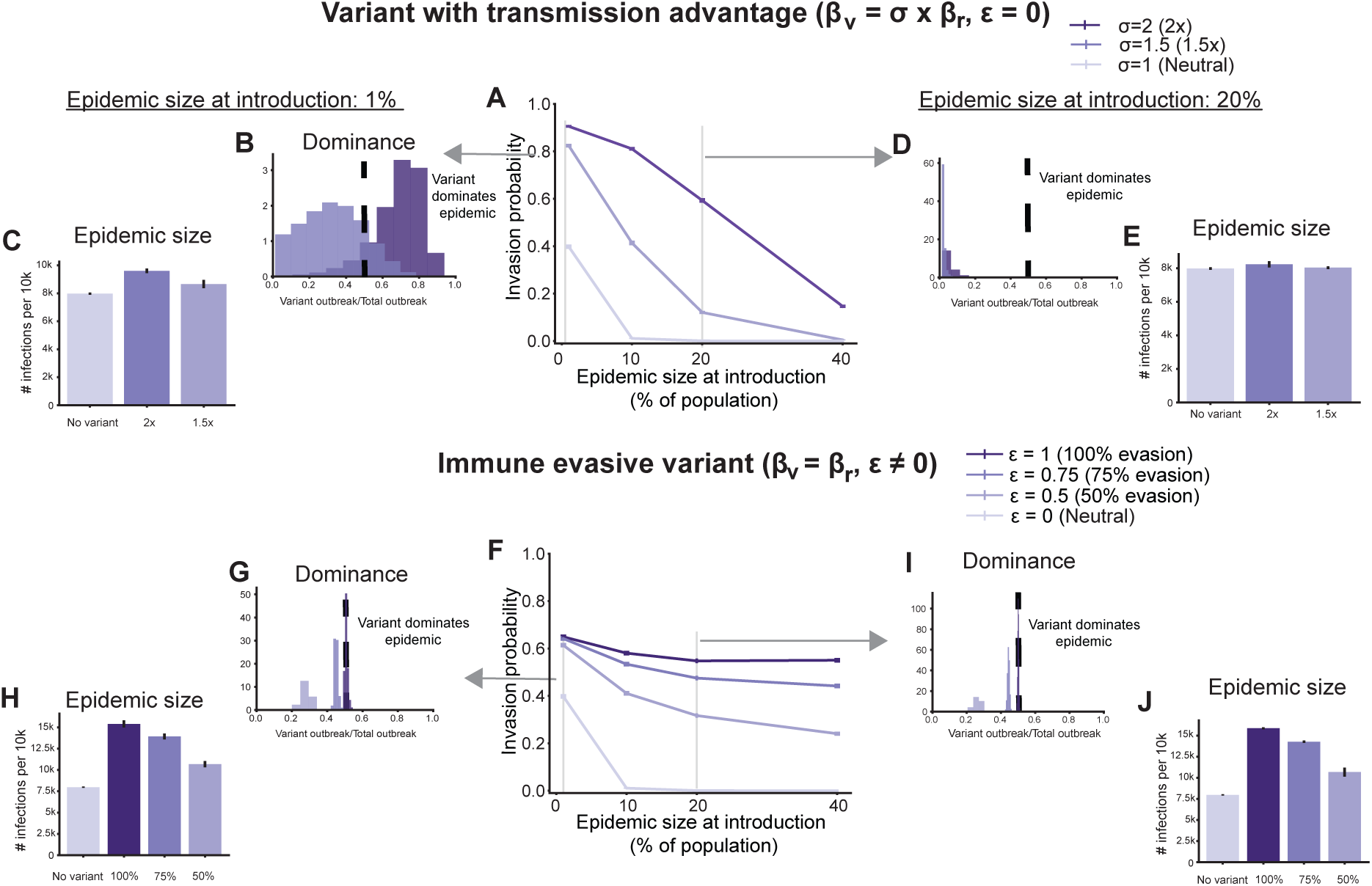
Effect of variant type and population immunity on variant invasion and epidemic impact when resident strain *R*_0_ = 2. Results of stochastic simulations for variants with transmission advantage (top row) and variants with immune evasive properties (bottom row). A)&F) Fraction of simulations where the variant infected more than 1% of the population as a function of the size of the resident strain epidemic at the time of introduction. The vertical lines are the standard error of proportion. Histograms of the relative size of the variant outbreak compared to the total epidemic size and the average total epidemic size for two introduction times: variant introduced when B)&C), G)&H) 1% and D)&E), I)&J) 20% (bottom) of the population has been infected by the resident strain. Darker colors correspond to variants with a higher advantage for both variant types. Results are for 10,000 iterations for each introduction time.

**Figure S4:**
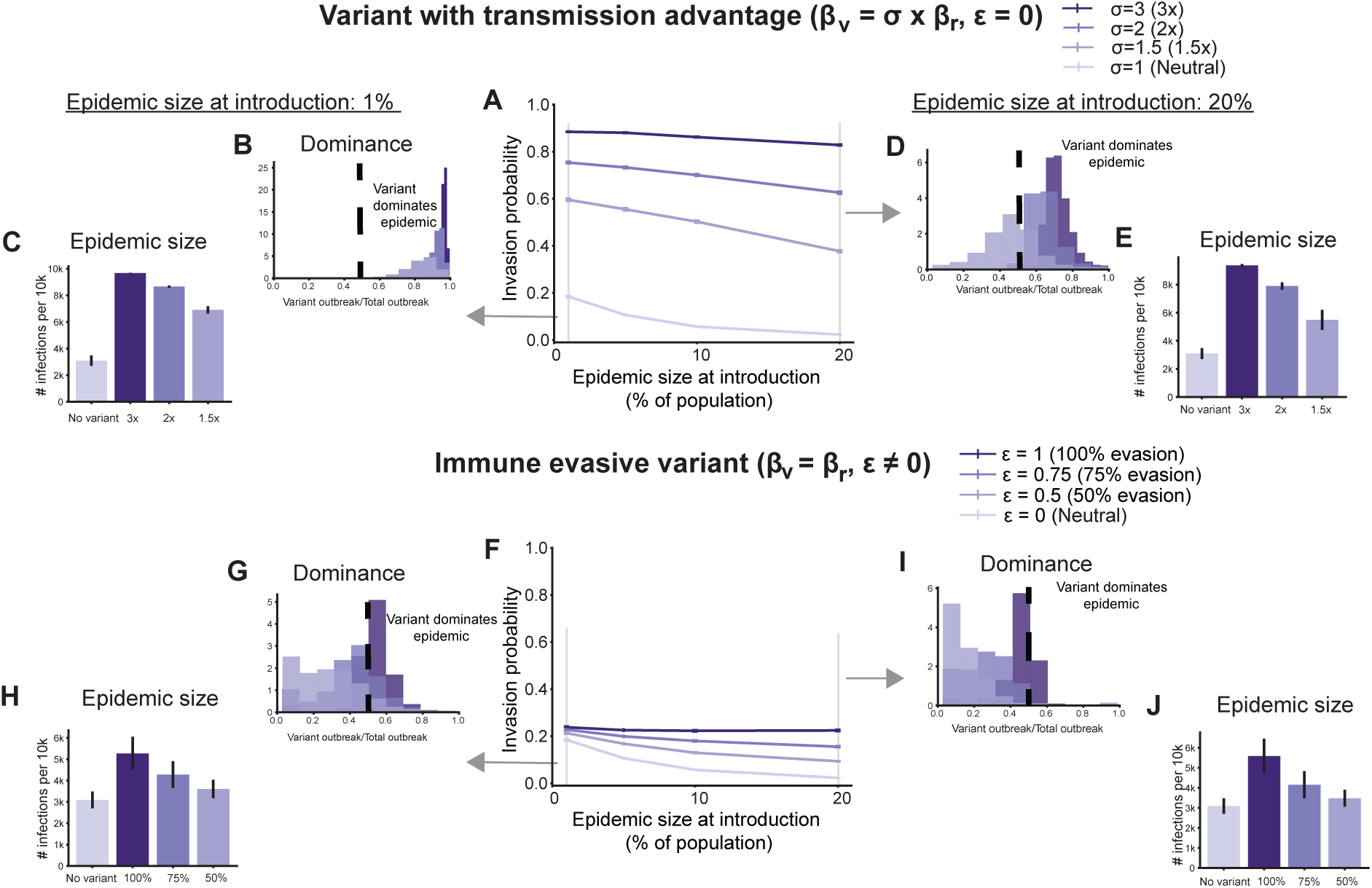
Effect of variant type and population immunity on variant invasion and epidemic impact when resident strain *R*_0_ = 1.2. Results of stochastic simulations for variants with transmission advantage (top row) and variants with immune evasive properties (bottom row). A)&F) Fraction of simulations where the variant infected more than 1% of the population as a function of the size of the resident strain epidemic at the time of introduction. The vertical lines are the standard error of proportion. Histograms of the relative size of the variant outbreak compared to the total epidemic size and the average total epidemic size for two introduction times: variant introduced when B)&C), G)&H) 1% and D)&E), I)&J) 20% (bottom) of the population has been infected by the resident strain. Darker colors correspond to variants with a higher advantage for both variant types. Results are for 10,000 iterations for each introduction time.

**Figure S5:**
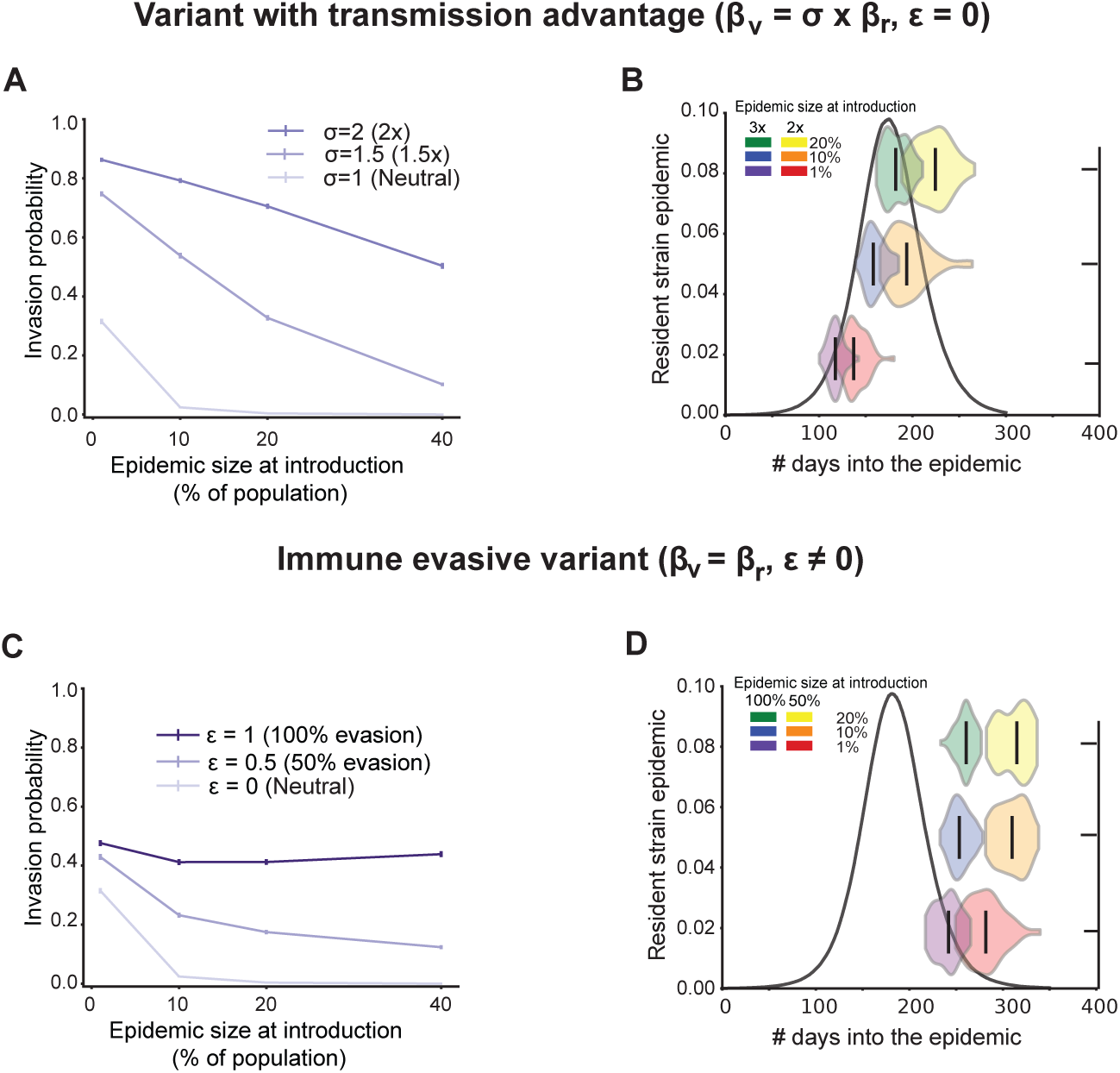
Invasion probability and detection times for variant under a longer duration of baseline infectiousness. Example results for a variant with transmission advantage (top row) and for an immune evasive variant (bottom row) with *R*_0_ = 1.5 and gamma-distributed duration of infectiousness with mean 14 and standard deviation of 7 days. A)& C) Fraction of simulations where the variant infected more than 1% of the population as a function of the size of the resident strain epidemic at the time of introduction. Darker colors correspond to variants with a higher advantage for both variant types. Results are for 10,000 iterations for each introduction time with the vertical lines denoting the standard error of proportion. B)&D) Violin plots of the time at which the variant reaches 5% prevalence in reference to the resident strain epidemic (black curves) for different introduction times. Results are for ∼ 50 iterations for each introduction time.

**Figure S6:**
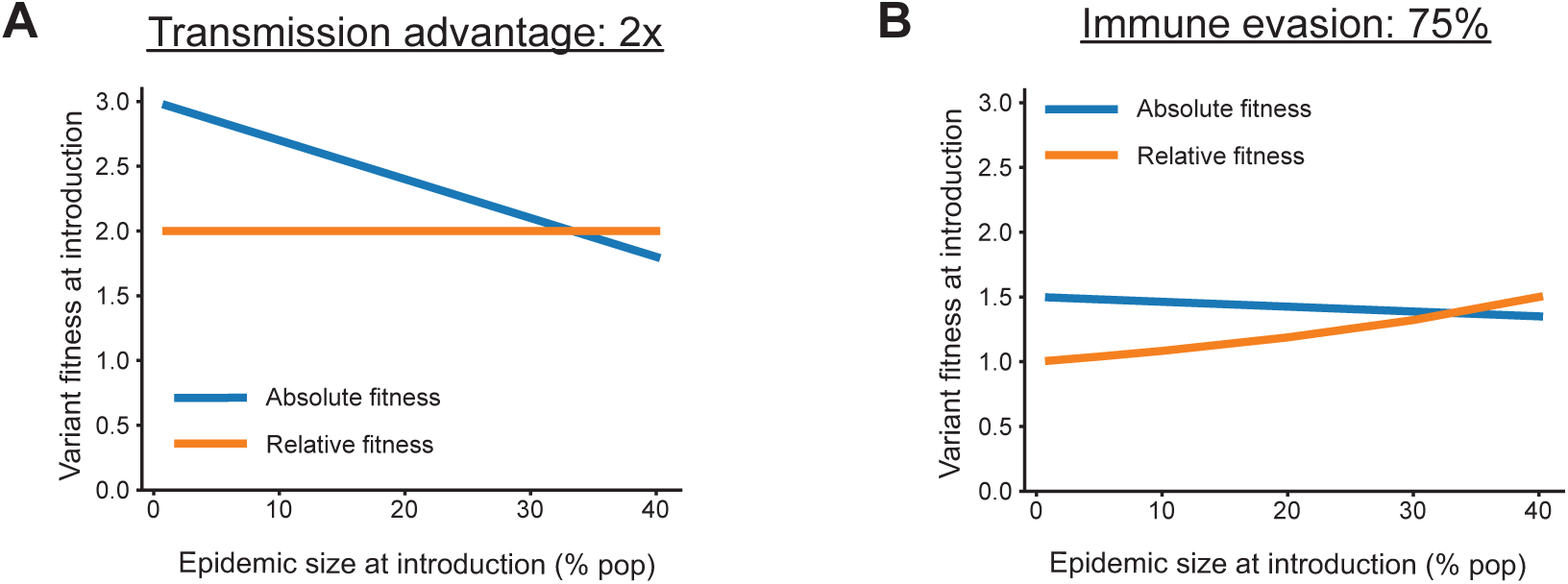
Example variant fitness at the time of introduction. The absolute and relative fitness of a variant as a function of the resident strain epidemic size at the time of its introduction into a resident strain epidemic when it is A) twice as transmissible as the resident strain and B) when it is 75% immune evasive. Resident strain *R*_0_ = 1.5

**Figure S7:**
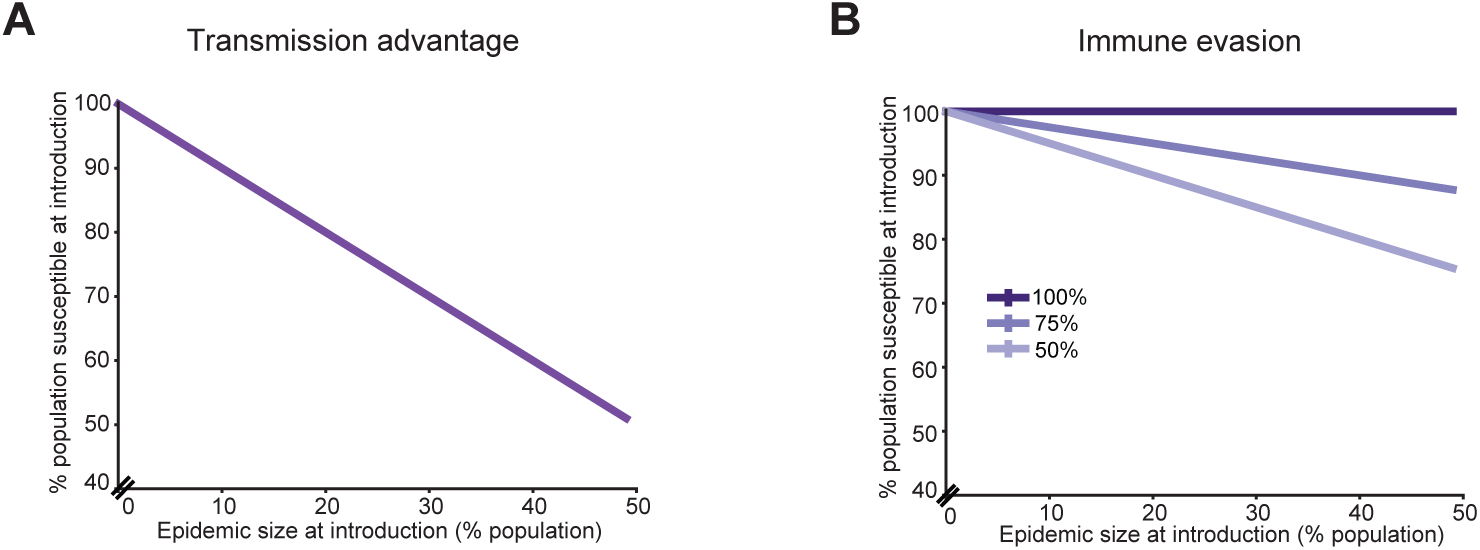
Amount of population susceptibility at the time of variant introduction. The percent of population that is susceptible to getting infected by the variant as a function of the resident strain epidemic size at the time of its introduction for A) a variant with transmission advantage and B) a variant with immune evasive properties. We provide a few levels of immune evasion as an example.

**Figure S8:**
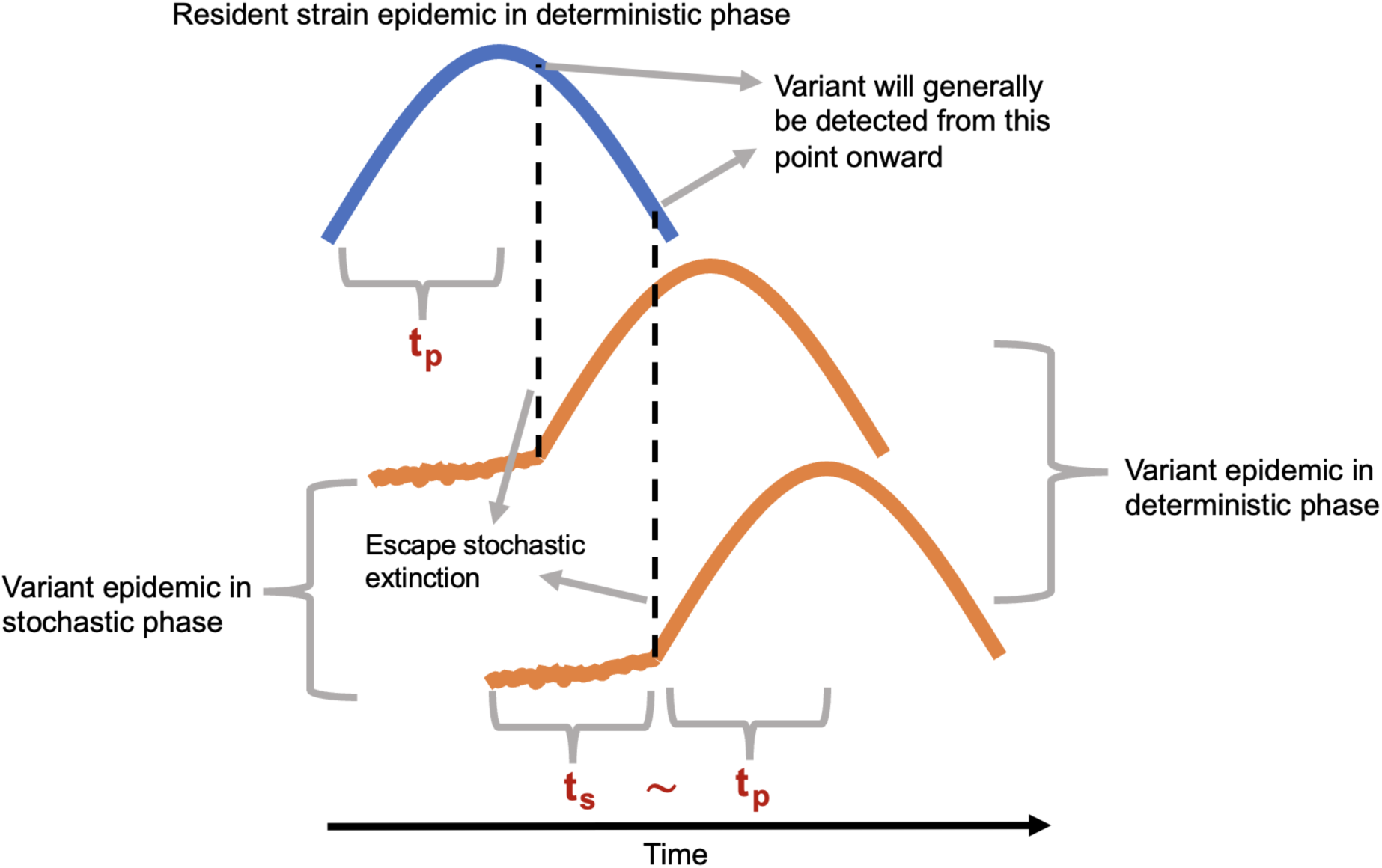
Schematic to provide intuition for time-scales in the variant invasion process. Blue curve is the schematic of an epidemic curve for the resident strain that is already in its deterministic phase of spread. Orange curves correspond to the variant that first undergoes a stochastic phase for time *t_s_* after which it enters in the deterministic phase once it escapes extinction. *t_p_* is the time taken from that point onward to the epidemic peak. The deterministic trajectories of the two strains are shown to be the the same as it is a schematic of a fully immune evasive variant with the same baseline *R*_0_. Using the birth-death process approximation we find *t_s_ ∼ t_p_*.

**Figure S9:**
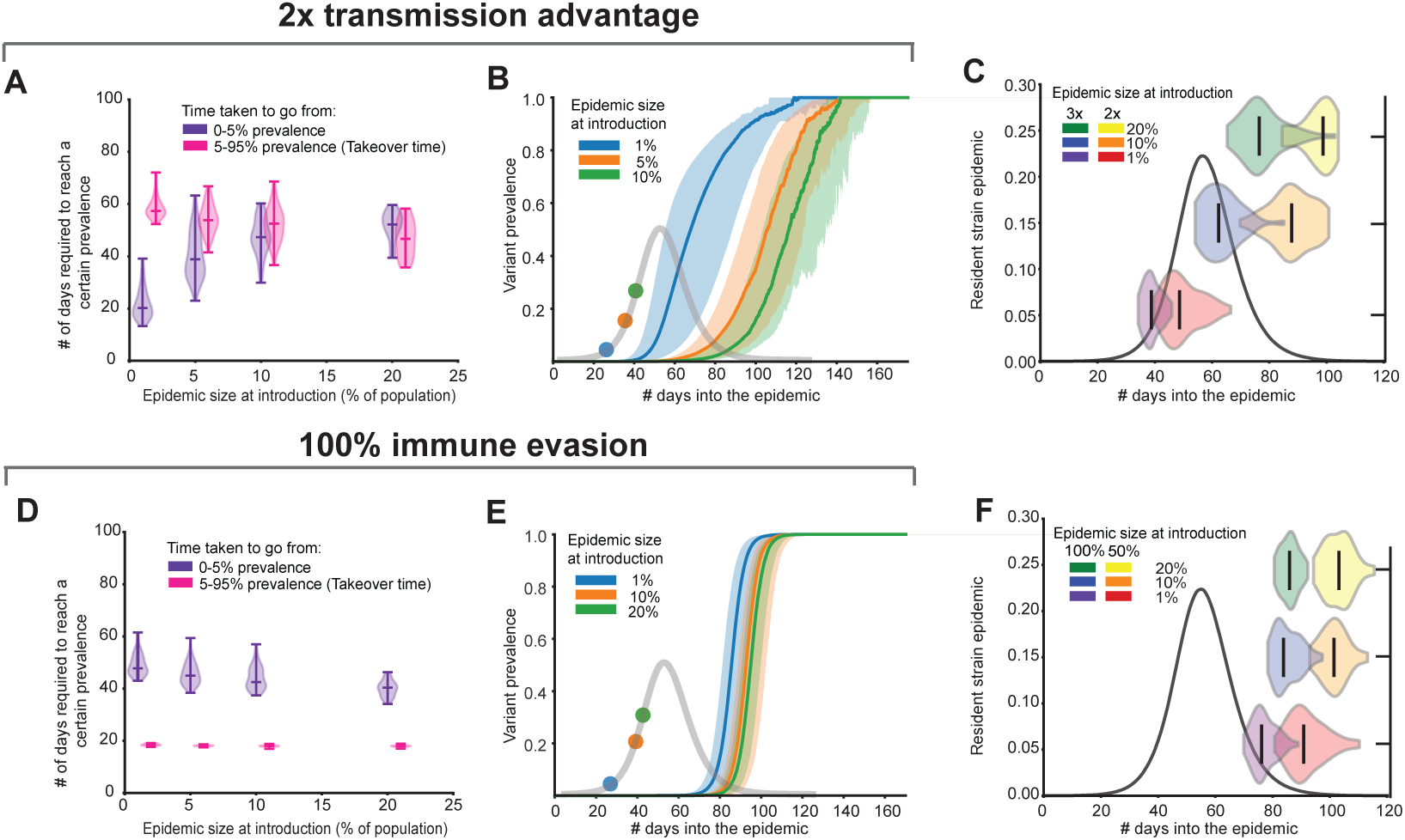
Rate of increase in variant prevalence as a function of the epidemic size at the time of introduction when resident strain *R*_0_ = 2. Example results for a variant with transmission advantage (top row) and for an immune evasive variant (bottom row) when the variant escapes stochastic extinction. A)&D) Number of days required for the variant to go from introduction to 5% and 5% – 95% prevalence as a function of the size of the resident strain epidemic at the time of introduction. B)&E) Variant prevalence over time when introduced at different levels of population immunity. Solid line is the median and the shaded region corresponds to the 5-95 percentile range. The resident strain epidemic in the absence of the variant is overlaid in light grey and marked with the respective introduction times. C)&F) Violin plots of the time at which the variant reaches 5% prevalence in reference to the resident strain epidemic (black curves) for different introduction times. Results are for 50 iterations for each introduction time.

**Figure S10:**
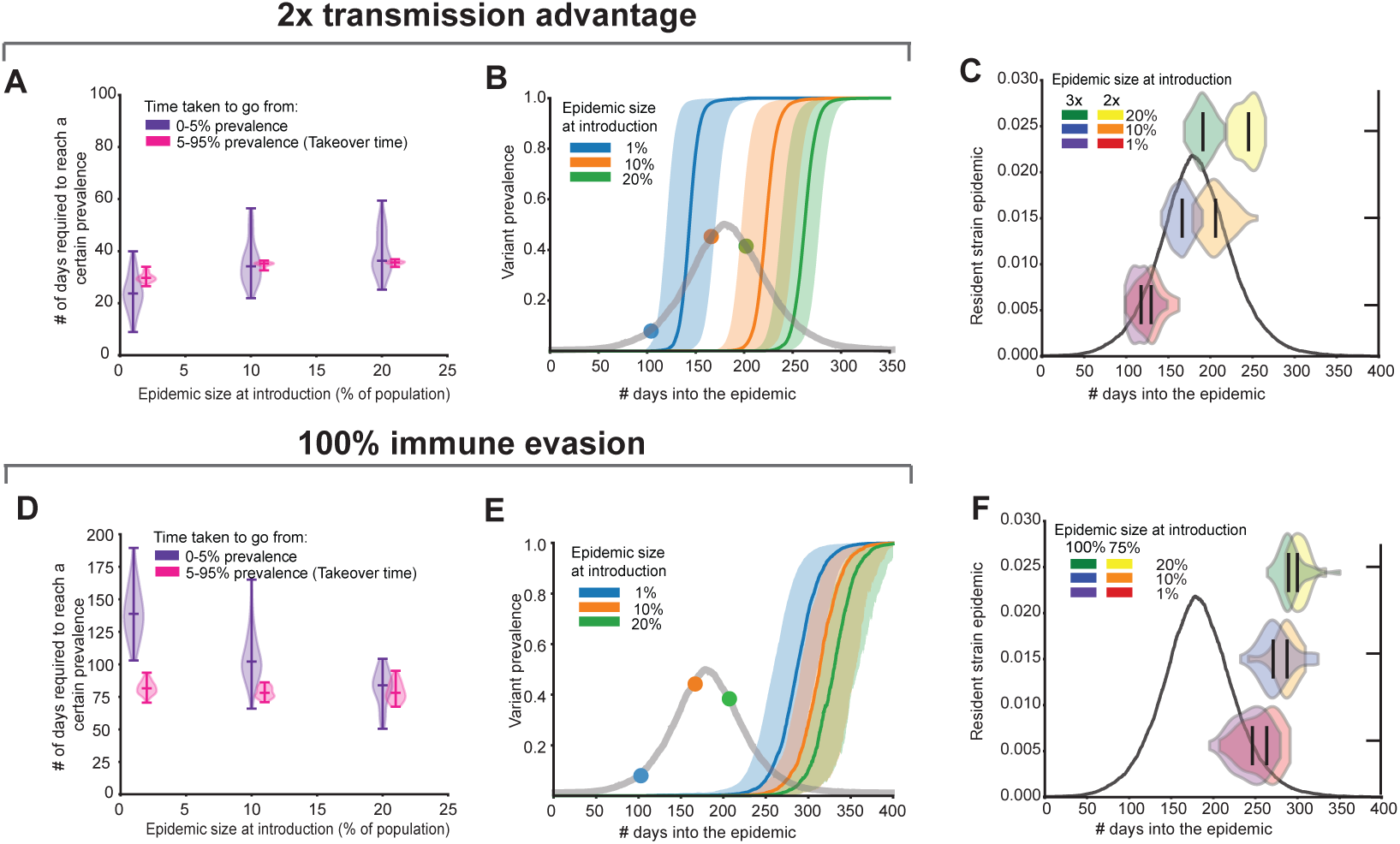
Rate of increase in variant prevalence as a function of the epidemic size at the time of introduction when resident strain *R*_0_ = 1.2. Example results for a variant with transmission advantage (top row) and for an immune evasive variant (bottom row) when the variant escapes stochastic extinction. A)&D) Number of days required for the variant to go from introduction to 5% and 5% – 95% prevalence as a function of the size of the resident strain epidemic at the time of introduction. B)&E) Variant prevalence over time when introduced at different levels of population immunity. Solid line is the median and the shaded region corresponds to the 5-95 percentile range. The resident strain epidemic in the absence of the variant is overlaid in light grey and marked with the respective introduction times. C)&F) Violin plots of the time at which the variant reaches 5% prevalence in reference to the resident strain epidemic (black curves) for different introduction times. Results are for ∼ 50 iterations for each introduction time.

**Figure S11:**
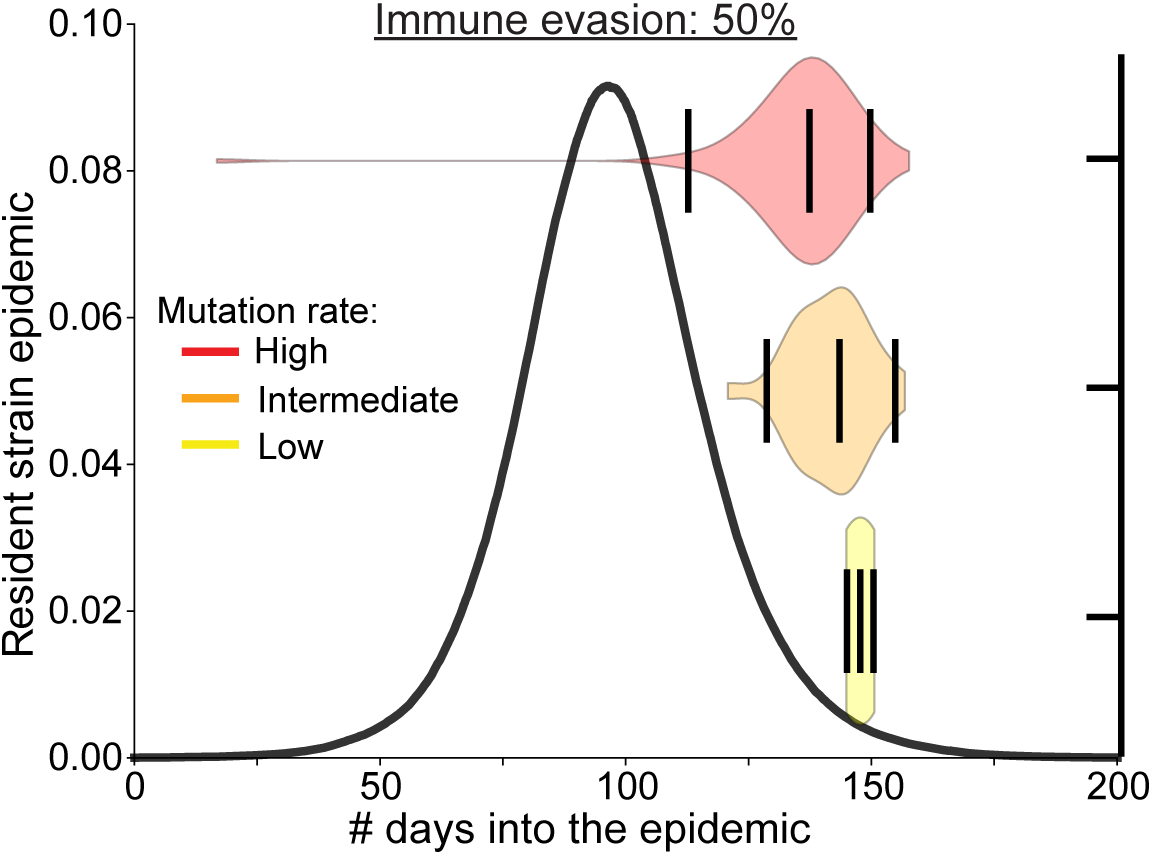
Variant detection times for different rates of evolution. Time at which a variant that is 50% immune evasive reaches 5% prevalence during the resident strain epidemic for different rates of evolution. The resident strain epidemic in the absence of the variant is provided as a reference (black curve). Results are for 100 iterations. The violin plots are marked with the median and the middle 95% quantile range.

**Figure S12:**
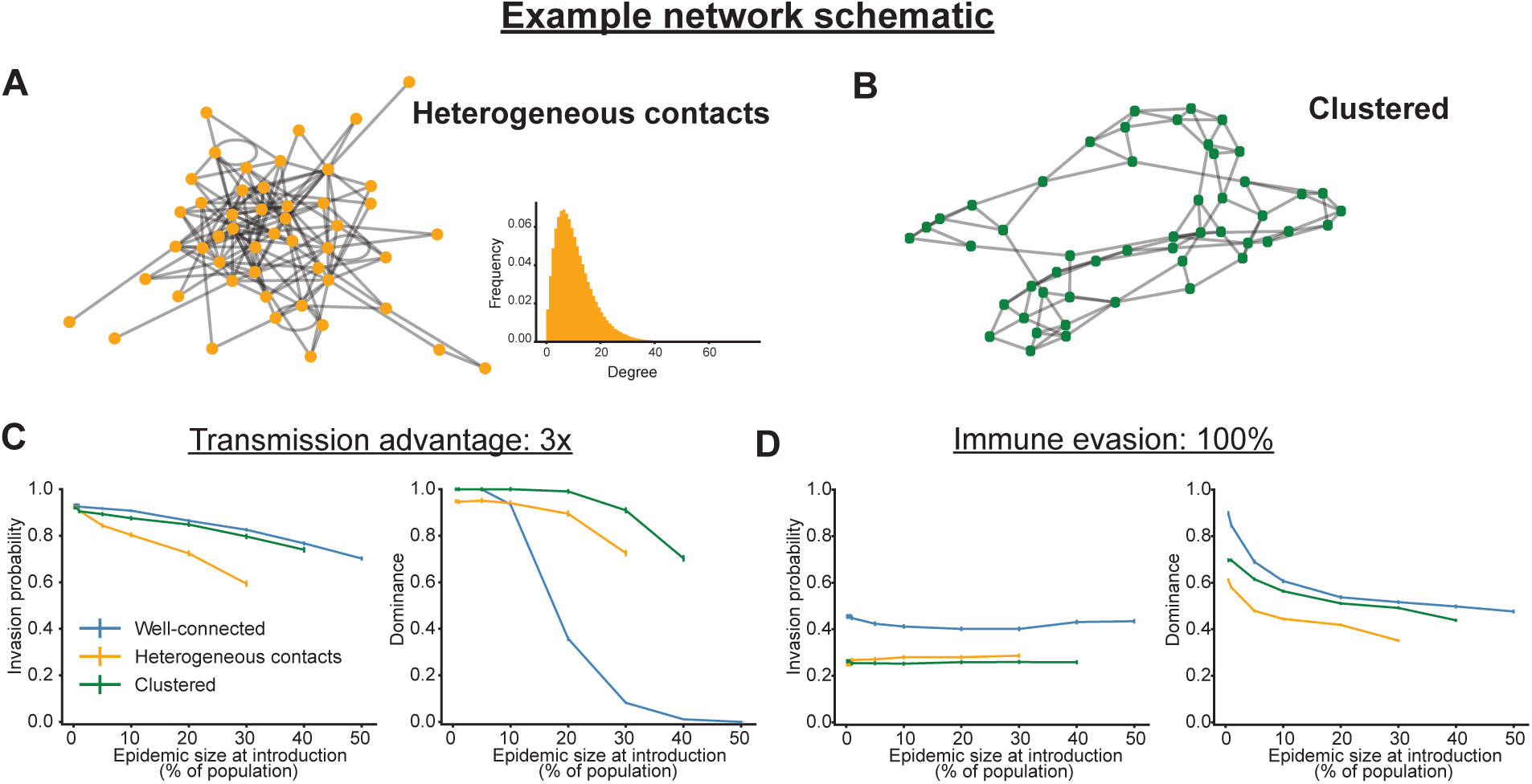
Schematic of the different types of networks considered and variant invasion and dominance results. A) Example of a network with heterogeneity in the number of contacts and the actual degree distribution used in this work. B) Example of a network with clustering of individuals where each individual as the same number of contacts. C)-D) Variant invasion probability and dominance for a C) transmission advantage and D) immune evasive variant for different underlying network structures. Invasion probability is defined as the fraction of simulations where the variant infected at least 1% of the population and dominance is the fraction of simulations where the variant dominated the epidemic. Results are for 5000 – 10, 000 iterations for each introduction time and error bars correspond to the standard error of proportion.

**Figure S13:**
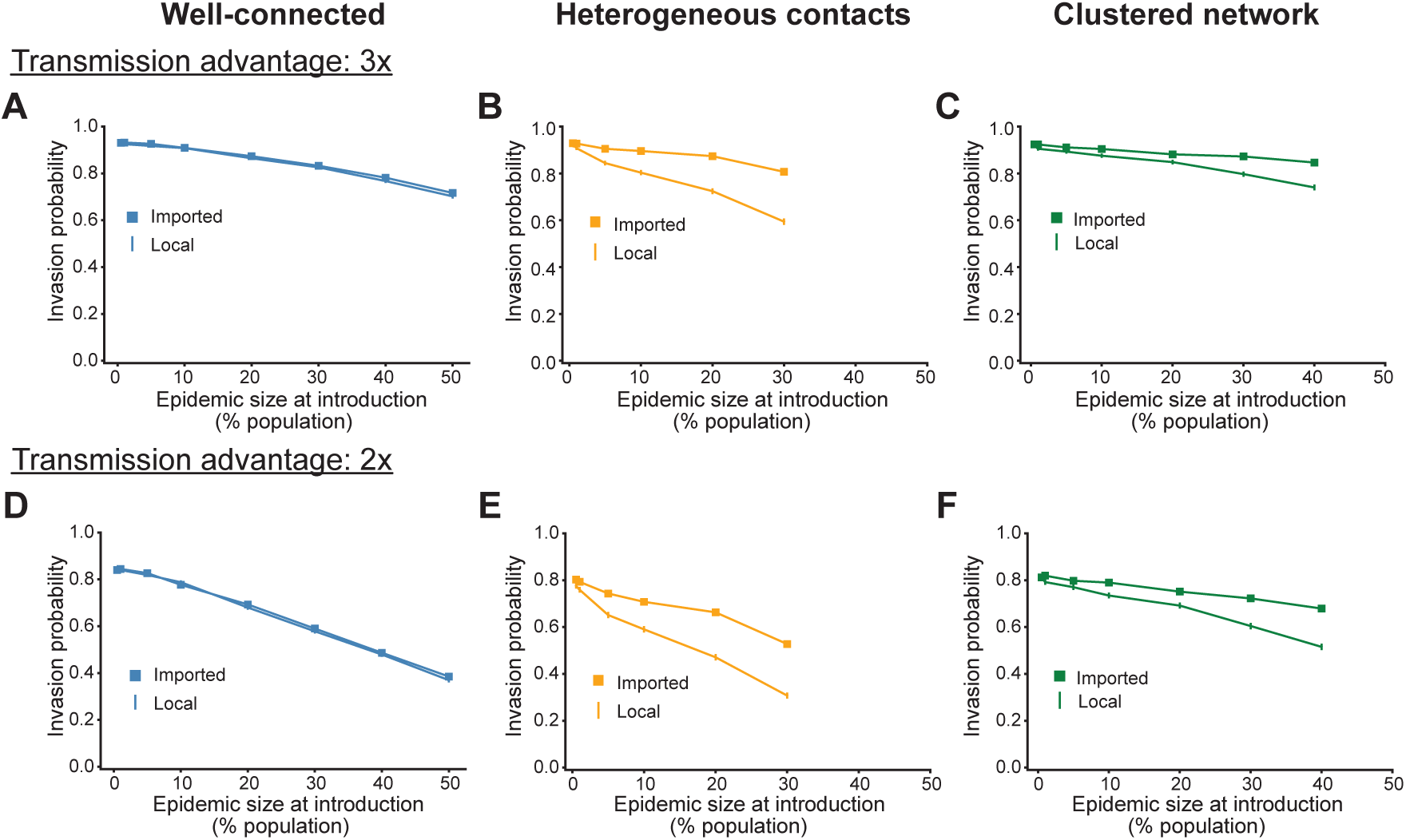
Invasion probability for two modes of variant introduction. Invasion probability of a variant as a function of the size of the epidemic when it is introduced for two example amounts of transmission advantage. Variant is either imported into (square) or evolves locally in the population (straight line) with different underlying structures. Results in subplots A)&D) are for a well-connected population, B)&E) for a population with heterogeneous number of contacts, and C)&F) clustered population. Invasion probability is defined as the fraction of simulations (n=10,000) where the variant infected at least 1% of the population. See SI Methods for network details.

**Figure S14:**
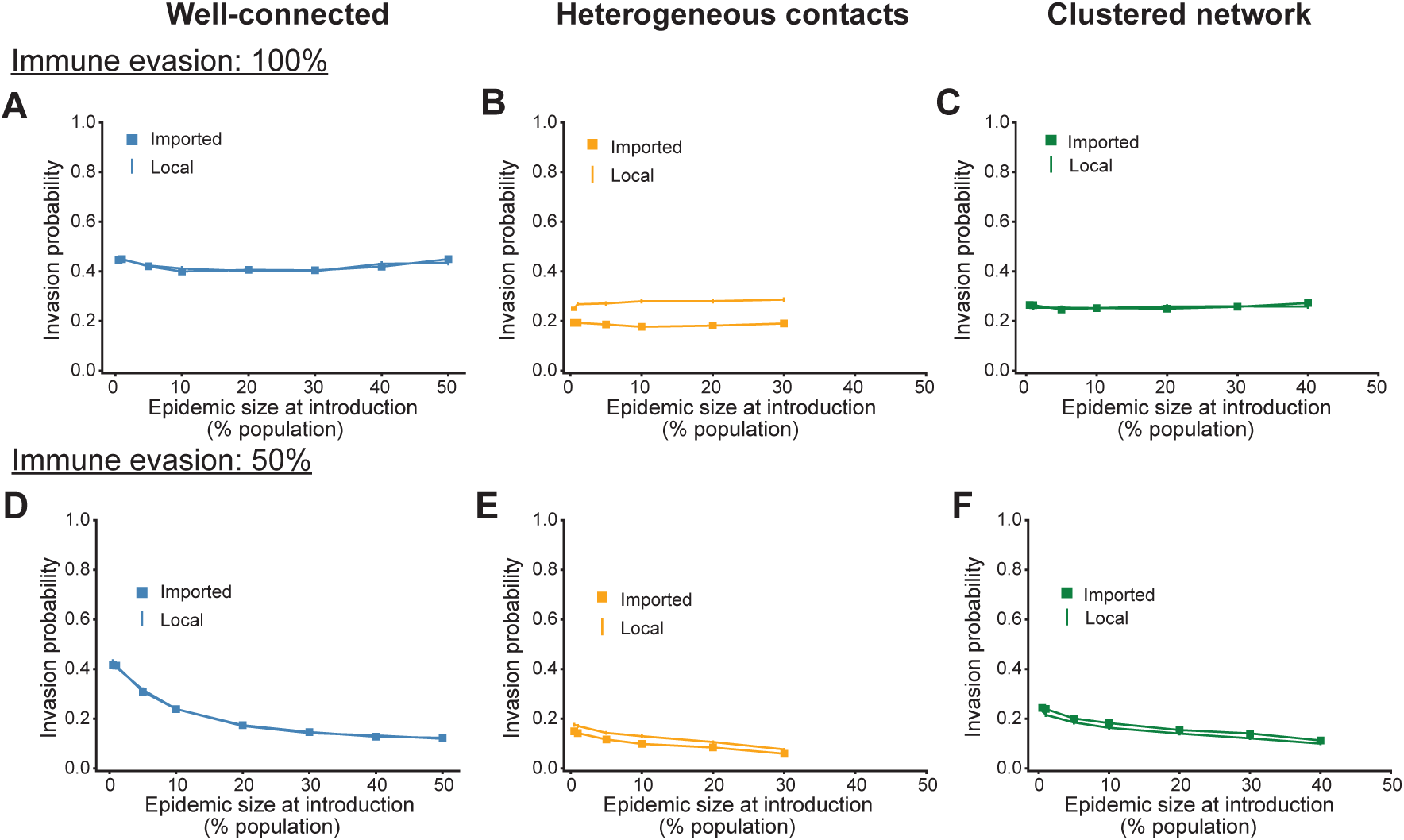
Invasion probability for two modes of variant introduction. Invasion probability of a variant as a function of the size of the epidemic when it is introduced for two example amounts of immune evasion. Variant is either imported into (square) or evolves locally in the population (straight line) with different underlying structures. Results in subplots A)&D) are for a well-connected population, B)&E) for a population with heterogeneous number of contacts, and C)&F) clustered population. Invasion probability is defined as the fraction of simulations (n=10,000) where the variant infected at least 1% of the population. See SI Methods for network details.

**Figure S15:**
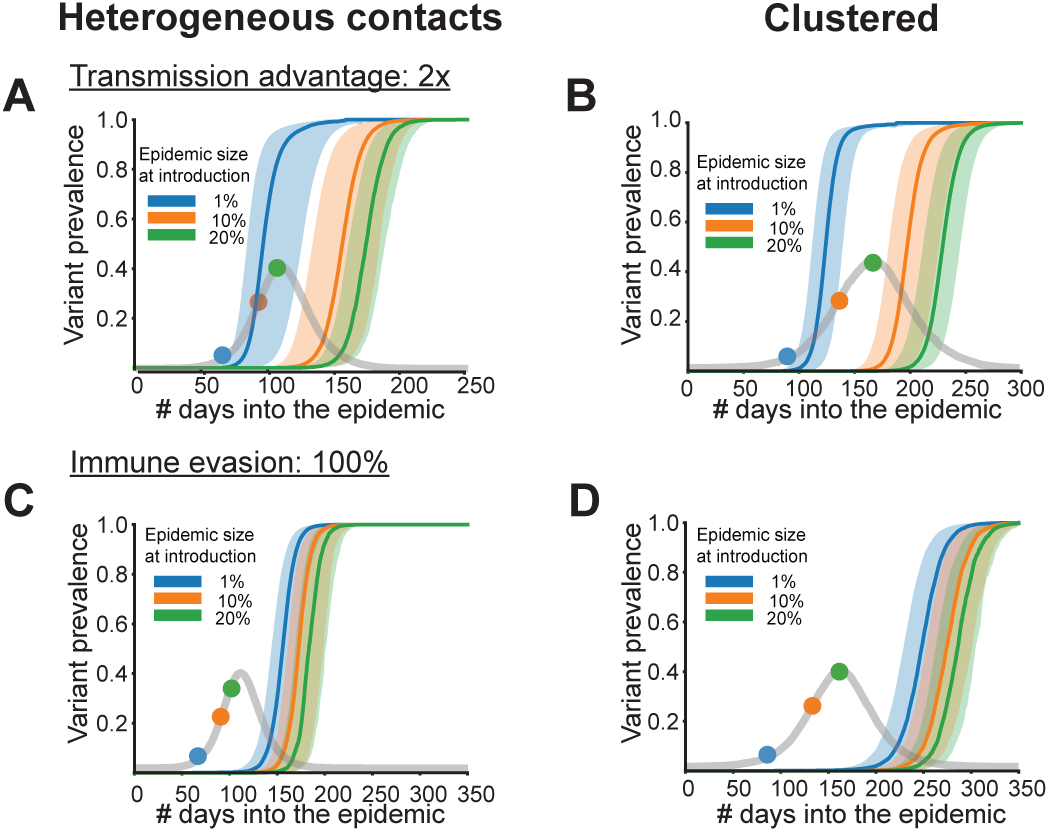
Dynamics of variant prevalence over time for the different underlying network structures. Variant prevalence as a function of time since the start of the resident strain epidemic for three example variant introduction times when the transmission network is either heterogeneous in the number of individual contacts (left) or is clustered (right). A)-B) Results are for a variant with a transmission advantage and C)-D) for an immune evasive variant. The resident strain epidemic in the absence of the variant is overlaid in grey along with the introduction times for reference. Results are for the baseline resident strain 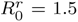 and for 50 iterations for each introduction time in a population of size 1 million. The solid line and shaded region corresponds to the median and the 5-95 percentile range respectively.

**Figure S16:**
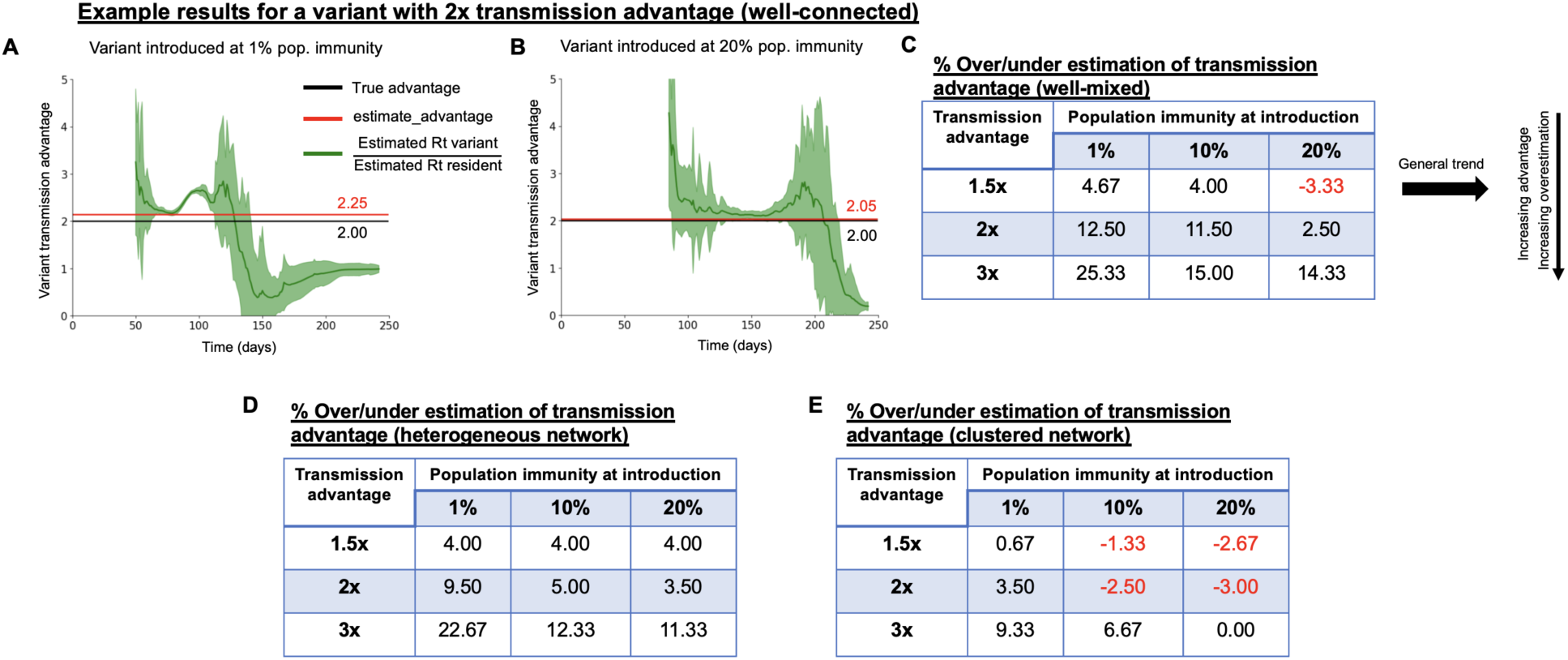
Ignoring competition can lead to an overestimation of the variant advantage. Estimated relative transmission advantage of the variant when it is introduced at A) 1% and B) 20% population immunity. Red line corresponds to the results obtained from the estimate_advantage function in EpiEstim, the green curve is obtained by taking the ratio of the variant and resident strain *R_t_*, and the black line corresponds to the true transmission advantage of the variant. C-E) Results of the estimate_advantage EpiEstim function for different levels of transmission advantage and introduction times for the well-connected, heterogeneous and clustered networks. Entries in red correspond to underestimation in the variant advantage. See Suppl. Analysis for more details.

**Figure S17:**
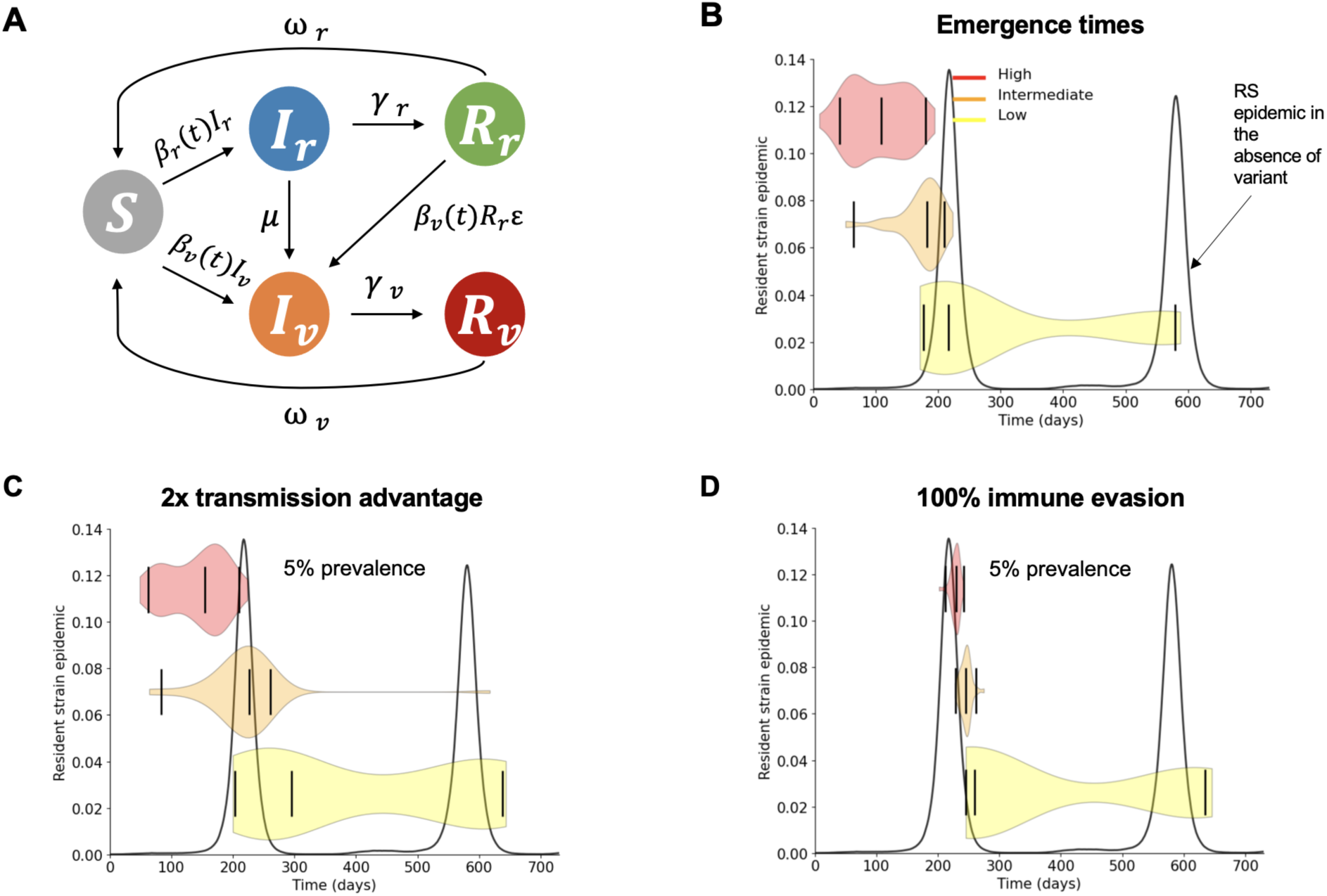
Model schematic and example variant detection times. A) Schematic of the two strain SIR-type model with a well-connected contact network consisting of individuals susceptible to infection (*S*), infected by the resident (*I_r_*) or variant (*I_v_*) strain, and recovered from infection by the resident (*R_r_*) or variant (*R_v_*) strain. For strain i, *β_i_* is the per contact transmission rate which is time dependent and *γ_i_* is the rate of recovery. Recovered individuals become susceptible at rate *ω_i_*. The variant can be produced via mutation (rate *µ*) of the resident strain or imported into the population from an outside source. Variants with immune evasive properties (0 *< ε <*= 1) can infect individuals recovered from the resident strain infection by a rate proportional to the strength of immune evasion *ε*. B) Variant emergence times for different rates of variant evolution. Times when the variant reaches 5% prevalence for a variant with C) transmission advantage and D) immune evasive properties. The resident strain epidemic in the absence of the variant is provided as a reference (black curve). Violin plots are marked with the median and the middle 95% quantile range. See Suppl. Analysis for more details.

